# Defining the potential impact and cost-effectiveness of a non-invasive diagnostic for malaria: a modeling study

**DOI:** 10.64898/2026.03.31.26349813

**Authors:** Megan A Hansen, Alexandra de Nooy, Serafina Calarco, Kevin KA Tetteh, Brooke E Nichols

**Author notes:** **Correspondence:** BEN and MAH.

## Abstract

**Background:** Malaria rapid diagnostic tests (RDTs) are widely used to detect and treat malaria infections, yet a diagnostic gap remains. With turnaround times of ∼15 minutes, RDTs may be too slow to enable broad-scale implementation in certain contexts. Novel non-invasive diagnostics (NIDs) have potential to provide faster (<5 minutes), sensitive (90% for symptomatic, 65% for asymptomatic carriage), and cost-effective alternatives, which may increase testing throughput, enhance case detection, guide appropriate antimicrobial use, and reduce waste by using fewer consumables. Their potential impact has yet to be investigated.

**Methods:** We modeled a country-agnostic population of 10 million individuals to assess the impact of population-level scale-up of four malaria testing strategies for active case-finding: 1) current practice (50% syndromic diagnosis and 50% RDTs), 2) full RDT scale-up, 3) full NID implementation, and 4) NID screening plus confirmatory RDT, using a decision-tree model of the malaria diagnostic and care cascade. We varied prevalence (0.02–0.25) and proportion of cases with symptoms (0.05–0.60) to evaluate strategy performance across epidemiological contexts. We investigated case detection rates, antimicrobial use, incremental cost-effectiveness ratios (ICERs) per disability adjusted life year (DALY) averted, net positive treatment outcomes, and threshold performance levels at which an NID would outperform RDTs.

**Results:** Full NID implementation (strategy 3) yielded the highest case detection rates (up to 85%), followed by strategies 2, 4, and 1 (45%, 38%, 36% respectively). NID-based methods (strategies 3 and 4) saved costs and RDT scale-up was cost-effective at averting DALYs compared to current practice (ICERs: $60–1,270). Despite high case detection, universal NID testing spiked unnecessary antimicrobial use. Overall, our results suggest that an NID with 55% asymptomatic sensitivity and 84% specificity, followed by RDT confirmation (strategy 4), could simultaneously improve case detection, reduce antimicrobial overuse, and limit costs.

**Conclusions:** This modeling analysis suggests that NIDs can sustainably optimize malaria case detection in symptomatic and asymptomatic cases and reduce costs, potentially making them a valuable addition to the diagnostic toolbox. When paired with confirmatory RDTs, they could help reduce inappropriate antimicrobial use, supporting drug efficacy amid rising resistance. Further research should assess their real-world utility, feasibility, and scalability for malaria surveillance and elimination efforts.

## Introduction

Malaria remains a major global health threat, with an estimated 263 million infections worldwide in 2023, primarily in Africa and Southeast Asia.^1^ Although substantial progress has been made over the past two decades in prevention, diagnosis, and treatment – including widespread use of artemisinin-based combination therapy (ACT) – malaria control and elimination efforts remain fragile and uneven.^1^ A central challenge is ensuring timely and accurate diagnosis for all individuals presenting for care, particularly in high-burden and resource-constrained settings. Effective surveillance and case management depend on timely and reliable diagnostic tools that perform well across a range of transmission settings and can be feasibly implemented at scale within existing health system constraints.^2^

Current malaria diagnostics include microscopy, rapid diagnostic tests (RDTs), and polymerase chain reaction (PCR).^3^ Microscopy remains the reference standard for parasite detection but requires skilled personnel and laboratory infrastructure, often resulting in delayed diagnosis.^4^ When a quick diagnosis is required, RDTs are used as a first-line approach to confirm or exclude malaria infection.^4^ Despite varying accuracy among these diagnostic tools, RDTs have been championed as cost-effective compared to microscopy in various transmission settings.^5,6^ However, full population coverage with RDT-based testing has not been achieved even in settings where RDTs are widely available, due to practical constraints that limit universal RDT uptake and consistent use. In high-volume clinics, the 15-minute turnaround time often exceeds staff capacity, especially during peak malaria seasons.^3^ Consequently, many febrile patients remain untested, leading to presumptive clinical diagnosis despite poor performance in differentiating between febrile illnesses.^7^ This diagnostic gap drives substantial antimalarial over-prescription, sometimes exacerbated by mass drug administration (MDA).^8,9^ Such excess use increases programmatic costs and threatens first-line therapies by accelerating the development and spread of drug resistance.^7,10^

Novel non-invasive diagnostic (NID) technologies have emerged as a potential complement or alternative to existing tests. NIDs do not require blood sampling and reduce reliance on trained personnel and laboratory infrastructure, representing a substantial shift from conventional malaria diagnostics.^11,12^ Recent landscape analyses by UNITAID and FIND identified multiple early-stage NID platforms that may detect low-density and asymptomatic infections more effectively than RDTs.^12–14^ These technologies generate results in seconds, require minimal consumables, and facilitate high-throughput screening in clinical and community settings.^14–18^ Such emerging NID methods enable rapid, painless malaria diagnosis by detecting parasite-induced physical, chemical, or optical changes in the host.^19^ Infrared spectroscopy analyzes tissue light absorption patterns to identify malaria-infected cells, often combined with computational algorithms to improve accuracy.^15^ Raman spectroscopy examines molecular vibrations in tissues to identify infected cells.^20^ Photoacoustic systems use laser pulses to generate acoustic waves that detect infected red blood cells based on unique optical and acoustic properties.^21,22^ Other approaches recognize malaria-associated volatile organic compounds in breath or skin emissions, reflecting parasite metabolism and enabling chemical biomarker-based diagnosis.^23,24^

However, the population-level impact, cost-effectiveness, and optimal role of NIDs in malaria diagnostic algorithms remain poorly defined, particularly in resource-limited and diverse epidemiological settings.^14,18^ It remains unclear how NIDs might address diagnostic gaps, reduce inappropriate prescribing, or improve health system efficiency relative to RDTs. We used a country-agnostic mathematical model to evaluate the impact and cost-effectiveness of NIDs, modeled as a technology class, against existing diagnostic strategies across varying prevalence scenarios. Beyond strategy comparison, we identify the minimum performance characteristics required for NIDs to become a value-adding intervention by achieving cost-effectiveness or improved antimicrobial stewardship relative to current practices and RDT testing alone.

## Methods

We used decision-tree modeling to compare the clinical and economic impact of four population-scale malaria testing strategies (**Figure 1**).

1) Current practice: a mix of syndromic diagnosis with RDTs (50%/50%)^9^
2) Full-scale RDT implementation: complete access to RDTs
3) Universal NID implementation: complete access to NIDs
4) NID screening with RDT confirmation: universal screening access to NIDs, followed by a confirmatory RDT for those who screen positive

**Figure 1.**
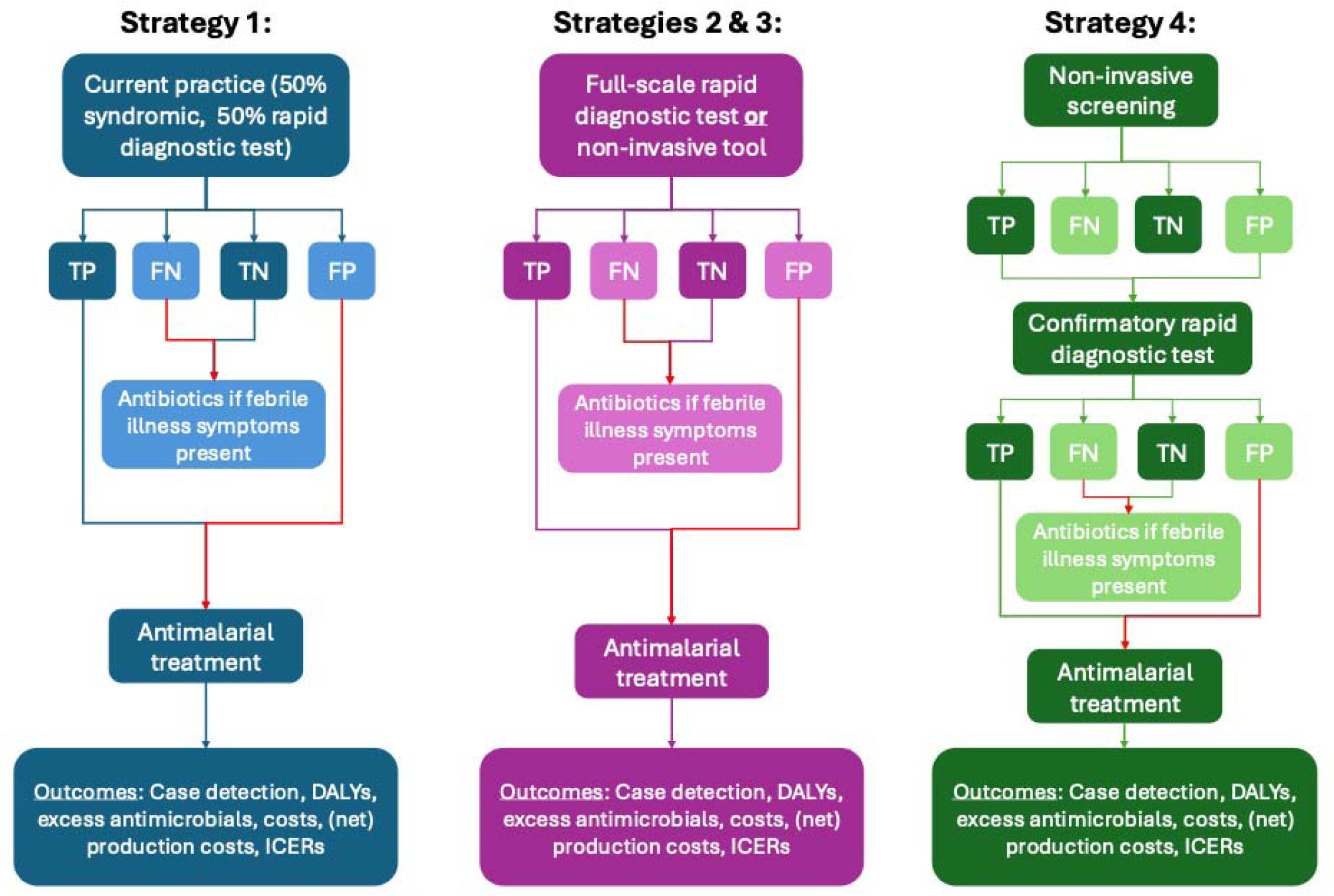
Conceptual diagram of the model. Abbreviations: TP: true-positive, FN: false-negative, TN: true-negative, FP: false-positive

### Model framework

The model simulated the malaria diagnosis and treatment cascade of a hypothetical country-agnostic cohort of 10 million individuals with a base-case malaria prevalence of 10% and a symptomatic proportion of 30%.^1,4^ We further varied prevalence (0.02–0.25) and the proportion of cases with symptoms (0.05–0.60) to evaluate strategy effectiveness under varying epidemiological conditions and burden levels.^2^ Model parameters were synthesized from published literature and population-level estimates to ensure generalizability across a range of malaria-endemic settings (**Table 1**). Modeling and analyses were completed in RStudio (Version 2023.12.1+402).

**Table 1.**
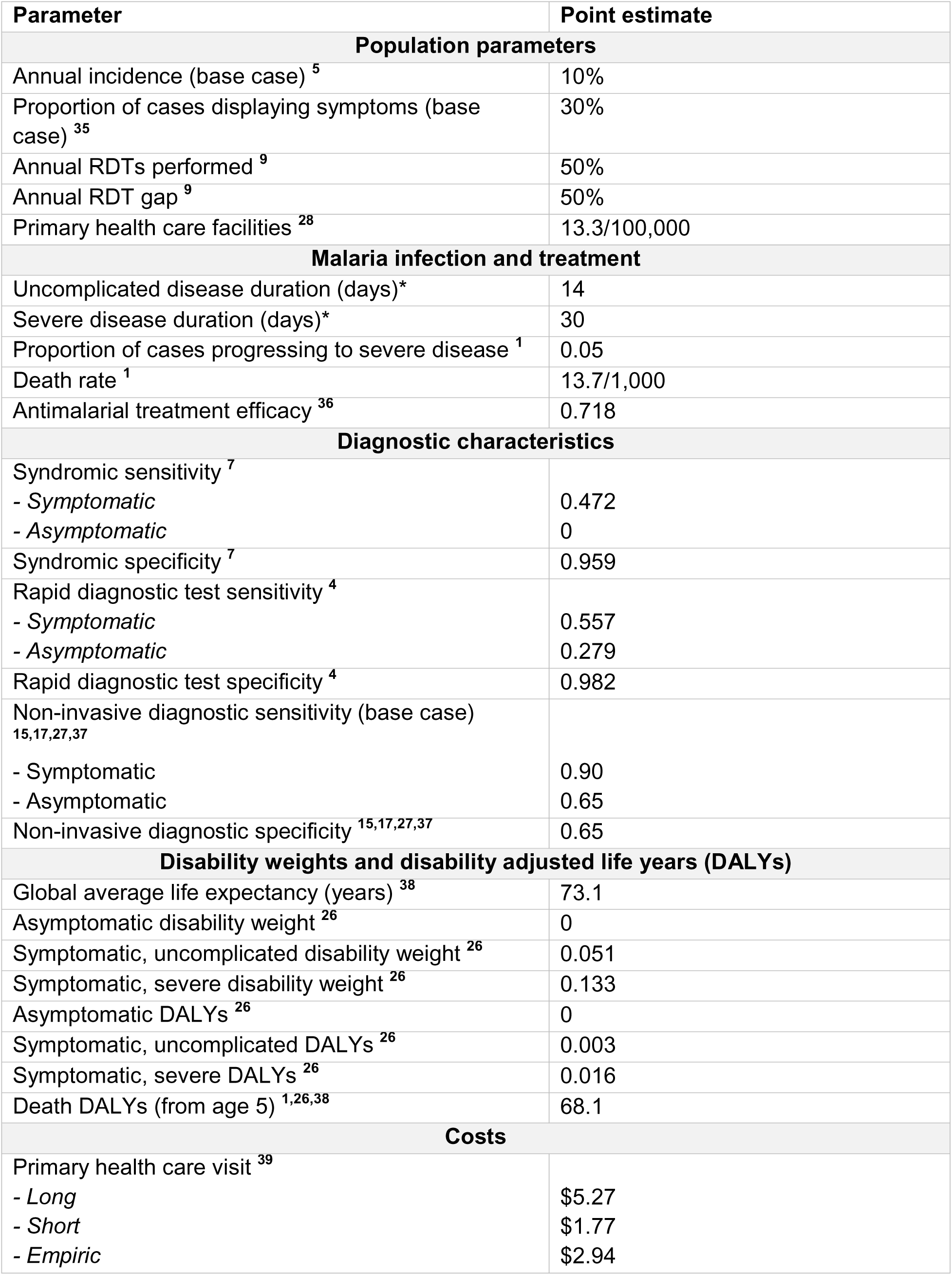
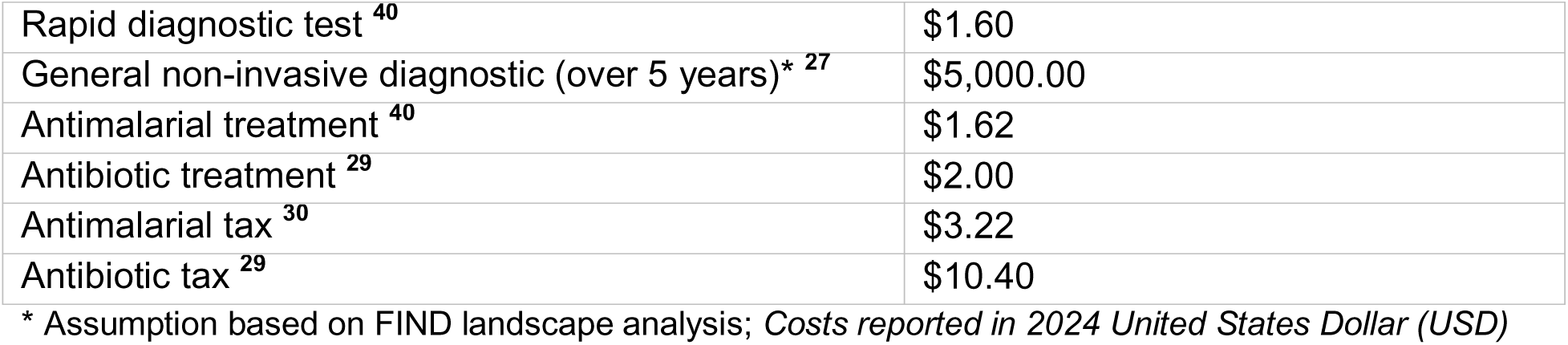
Parameters and costs for model input.

The decision-tree model determined the diagnostic yield (i.e., true/false-positives/negatives) across all modeled prevalence and symptomatic levels. Following the diagnostic cascade, ACT was prescribed to individuals with positive results regardless of symptom status and whether the diagnosis was correct. Symptomatic patients with negative results were prescribed amoxicillin; these were classified as inappropriate and ineffective treatments contributing antimicrobial resistance when given to individuals with a false-negative test.^25^ No treatment was administered to asymptomatic cases with negative results.

Disease burden was modeled based on disability weights and duration of illness for uncomplicated and severe malaria, with a malaria-specific mortality rate of 13.7 per 1,000 (**Table 1**).^1,2,26^ DALYs were calculated as the sum of years of life lost (YLL) due to premature mortality and years lived with disability (YLD) from morbidity. YLL were computed using the WHO average global life expectancy, and YLD were derived using disability weights from the IHME 2021 Global Burden of Disease Study.^26^ DALYs averted were the difference between baseline DALYs (i.e., current practice) and the DALYs averted with each alternative testing strategy.

### Economic modeling and cost-effectiveness analysis

Diagnostic costs were assessed from a provider perspective, including the cost of a primary health care (PHC) visit, RDTs, and NIDs. Unit costs incorporated healthcare worker time, facility-level implementation costs across PHC clinics, and an expected 5-year working life of many NIDs (**Table 1**).^27,28^. This analysis assumes a consumable-free, instrument-based NID, with one instrument deployed per healthcare clinic. Since expected testing throughput directly affects the cost per test, we estimated the distribution of healthcare clinics across the modeled population. Therefore, clinic density was standardized at 13.3 per 100,000 population, corresponding to the World Health Organization African Region average reported by the Global Health Observatory Data Repository.^28^

Treatment costs included ACT, amoxicillin, and ’AMR taxes’ to account for the long-term societal burden of inappropriate prescribing.^29^ The tax of an inappropriate antimalarial was considered to be the sum of an additional RDT plus one course of artemisinin.^30^ The antibiotic tax was derived from the long-term costs of one unnecessary broad-spectrum penicillin course, or amoxicillin.^29^ Further cost details for the four testing strategies are provided in **Text S1**. All costs were adjusted to 2024 US dollars (USD).^31^ We performed a cost-effectiveness analysis over a one-year horizon to assess the impact of various testing strategies on disease burden. Incremental cost-effectiveness ratios (ICERs) were calculated as the difference in total costs divided by the difference in DALYs averted for each epidemiological scenario. This enabled the identification of strategies defining the cost-effectiveness frontier. Study reporting adhered to the Consolidated Health Economic Evaluation Reporting Standards (**Table S1**).^32^

In a secondary cost analysis, we propose a new metric, the ‘cost per net correct diagnosis’. This is similar to production costs (i.e., cost per person correctly treated); however, the cost per net correct diagnosis considers both cases correctly identified and unnecessary treatment due to incorrect diagnoses given the real and present threat of AMR and concern for antimicrobial overuse (**Equation 1**).^33,34^ This new metric underscores that a diagnostic should fundamentally aim to treat more people correctly than incorrectly. In a threshold analysis, we determined the minimum NID specificity required across all modeled epidemic conditions to yield a positive cost per net correct diagnosis, defined as identifying more true-positives than incorrectly treated individuals. To highlight financial drivers of downstream resistance, we quantified the proportion of total costs attributable to inappropriate antimicrobial consumption.

### Sensitivity analysis

As NID technical specifications are not yet established, we further varied modelled asymptomatic sensitivity from 27.9% (RDT-equivalent) to 55.0% (nearly two-fold increase) for Strategies 3 and 4. Outcomes for these strategies were directly compared with full-scale RDT implementation, allowing us to assess the incremental clinical utility of enhanced diagnostic sensitivity for asymptomatic carriers. This enabled a comparison of NID and RDT performance for outcomes related specifically to asymptomatic infections, allowing us to identify the operational contexts and minimum test performance thresholds at which NIDs would provide greater utility than RDTs alone.

To evaluate the robustness of strategy performance to key epidemiological assumptions, we conducted a sensitivity analysis varying both prevalence and symptomatic proportion. Parameter combinations were grouped into five epidemic archetypes representing distinct epidemiological scenarios for outcome evaluation:

1) Base case: medium prevalence (0.10) medium symptomatic proportion (0.30)
2) Low prevalence (0.02), low symptomatic proportion (0.10)
3) High prevalence (0.20), low symptomatic proportion (0.05)
4) Low prevalence (0.05), high symptomatic proportion (0.60)
5) High prevalence (0.25), high symptomatic proportion (0.40)

We then compared outcomes across these epidemiologic conditions to assess how relationships between outcomes shifted across contexts relative to the base case (**Supplementary Appendix**).

**Equation 1**. Cost per net correct diagnosis.

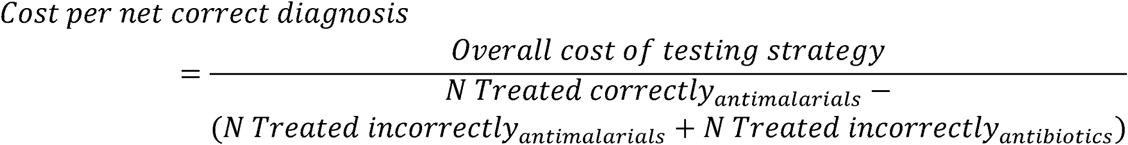

## Results

### At baseline diagnostic accuracy, NIDs improve case detection and reduce costs versus current practice and full RDT use, but have drawbacks

When comparing across testing strategies and using the baseline test accuracies described in **Table 1**, universal NID implementation yielded the highest case detection rates (up to 65%), followed by full-scale RDT use (up to 45%), current practice (up to 36%), and lastly NID screening followed by RDT confirmation (up to 29%) (**Figure S1A**). Universal NID testing also resulted in the greatest amount of true-positive results (470,000), but the greatest amount of false-positive results (3.15 million) in the base case scenario, primarily driven by the high sensitivity and assumed low specificity of NIDs (**Figure S1B**).^2,35^ Universal NID implementation also averted the most DALYs (57,890–1.52 million) at the lowest costs ($30–650 per DALY averted). Full-scale RDT use resulted in the next most DALYs averted (54,670–1.04 million) accompanied by a cost of $90–1,320 per DALY averted (**Figure S2**).

Despite universal NID testing yielding the lowest production costs, it consistently generated negative net production costs (i.e., more excess antimicrobials than true-positive results) (**Figure S3**). This is likely attributable to excessive antimalarials prescribed to individuals falsely testing positive with the NID; excess antimalarials spiked 2.63–3.43 million with universal NID testing as a sole diagnostic tool (**Figure S4**). Up to 49% of the costs associated with universal NID testing were attributable to unnecessary prescriptions (**Figure S5**). Although AMR accounted for a substantial share of total costs under the universal NID testing strategy, this approach remained cost-saving across all modeled epidemic conditions when compared with current practice (**Figure S6**). NID testing yielded the lowest projected long-term (5-year) costs among all testing strategies (up to $129 million) (**Figure S7**).

### At baseline diagnostic accuracies, NID screening followed by RDT confirmation can fill gaps to support appropriate treatment and remains competitive with current practice and full-scale RDT use

NID screening followed by a confirmatory RDT for those who screen positive, resulted in the greatest number of true-negative results (up to 8.94 million) when modeled at baseline accuracies, attributable to high RDT specificity (98%) and its ability to parse out individuals with a false-positive NID screening result (**Figure S1B**). However, it was often outperformed by all other strategies in case-detection rates, DALYs averted, and cost per DALY averted (**Figures S1A, S2**). NID screening followed by a confirmatory RDT resulted in similar production costs to current practice and full-scale RDT ($90–2,720), and most often generated positive net production costs of lower value (i.e., more true-positive results than excess antimicrobials prescribed, achieved at lower costs), due to low amounts of overall unnecessary antimicrobials (47,200–432,990) (**Figures S3, S4**). Threshold analysis shows that 65% NID specificity is adequate for most settings, but this requirement increases to 84% when prevalence falls below 5% to maintain a positive net cost per correct diagnosis (**Figure S8**).

NID screening with RDT confirmation achieved the highest level of antimicrobial stewardship among testing strategies, leading to the lowest share of costs attributable to unnecessary prescriptions (maximum 7%) (**Figure S4, S5**). While NID screening with a confirmation RDT was competitive with full-scale RDT implementation on many fronts, it was cost-saving relative to current-practice, unlike full-scale RDT use which yielded ICERs ranging $60–1,270 (**Figure S6**). NID screening with confirmatory RDTs resulted in lower long-term costs than full-scale RDT use ($271 million; only 76% of RDT costs) with only an 8% higher budget than current practice (**Figure S7**).

### Directly comparing NIDs to RDTs as a sensitivity analysis offers insights for improved diagnostic accuracy

When asymptomatic sensitivity of the NID matched that of the RDT (27.9%), it outperformed the RDT in case detection when symptomatic sensitivity was 60% or higher across all test specificities. Increasing NID asymptomatic sensitivity to 55% yielded the highest case detection rates (up to 68%), consistently outperforming full-scale RDT implementation. In contrast, NID with confirmatory RDT was uniformly outperformed by full-scale RDTs in case detection (**Figure 2A).** These gains came with increased antimicrobial use as universal NID implementation produced more unnecessary prescriptions than RDTs alone except at modeled 99% specificity (**Figure 2B**). NID screening with confirmatory RDTs was the most conservative strategy, producing no greater excess antimicrobial use than RDTs alone (**Figure 2B**).

**Figure 2.**
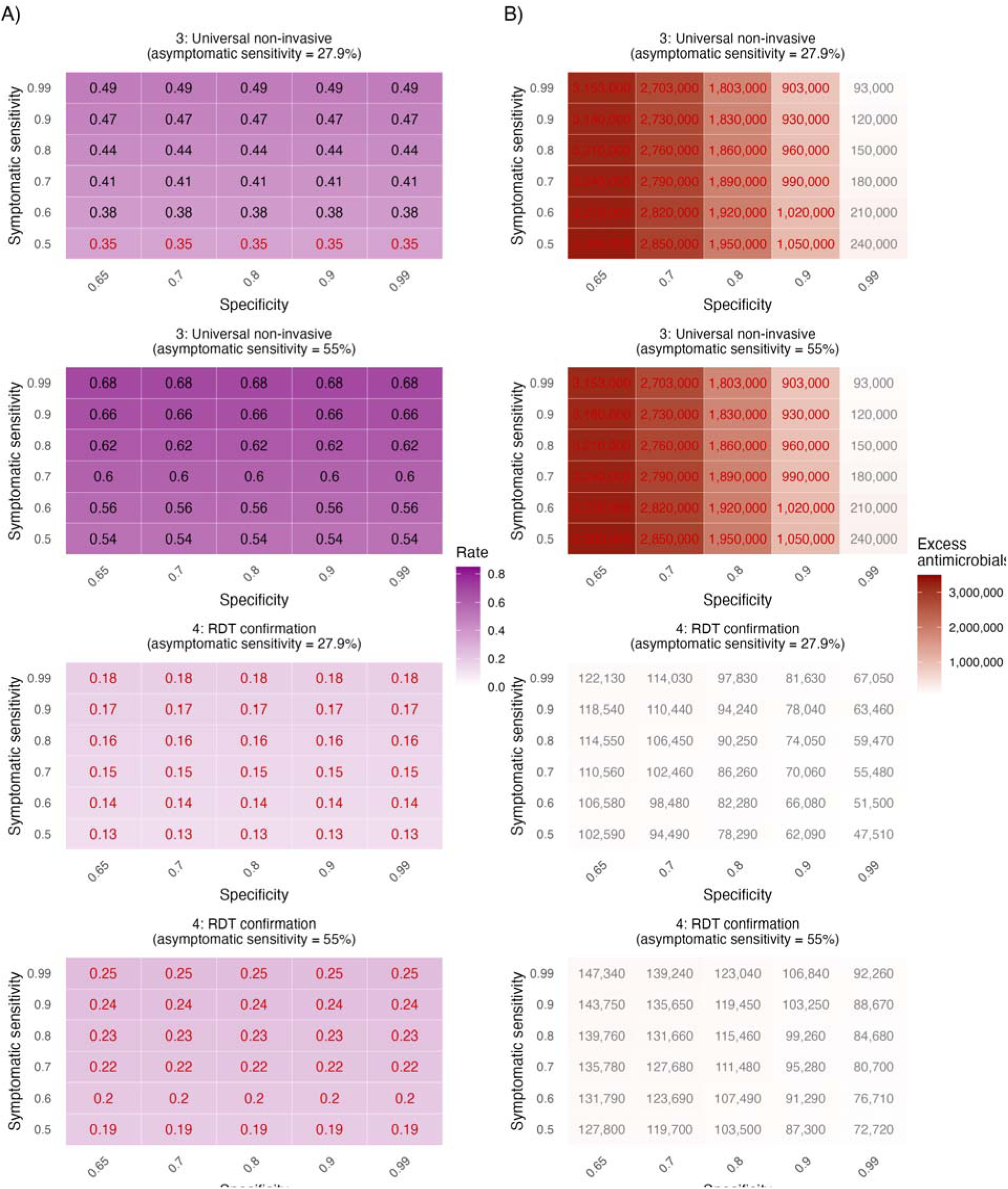
**A)** Case detection rates for universal non-invasive diagnostic implementation and non-invasive screening with rapid diagnostic test (RDT) confirmation (strategies 3 and 4) with varied asymptomatic sensitivity (27.9% and 55%). Results are shown for base case scenario. Darker violet tiles indicate higher case detection rates; red text indicates performance lower than a full RDT scale-up. **B)** Excess prescriptions (antimalarials and antimicrobials) for universal non-invasive diagnostic implementation and non-invasive screening with rapid diagnostic test (RDT) confirmation (strategies 3 and 4) with varied asymptomatic sensitivity (27.9% and 55%). Results are shown for base case scenario. Darker red tiles indicate higher excess prescriptions; red text indicates performance lower than a full RDT scale-up.

DALYs averted closely mirrored case detection rates, given their direct link (**Figure 3A**). At a modeled asymptomatic sensitivity of 27.9%, the NID averted between 322,700 and 460,390 DALYs – exceeding full-scale RDT use except when symptomatic sensitivity was set to 50%. This performance was achieved at a cost of approximately $60–130 per DALY across modeled scenarios (**Figure 3B**). NID screening with confirmatory RDT testing averted substantially fewer DALYs at the modeled asymptomatic sensitivity of 27.9% and was consistently outperformed by full-scale RDT use (**Figure 3A**). The cost per DALY averted ranged $160–370, also making it outperformed by full-scale RDT implementation except with higher symptomatic sensitivity and specificity (e.g., 80%) (**Figure 3B**). Since zero DALYs are assigned to asymptomatic cases in the model, there was no change in DALYs averted when doubling the NID’s asymptomatic sensitivity, thus results are not shown in **Figure 3**.

**Figure 3.**
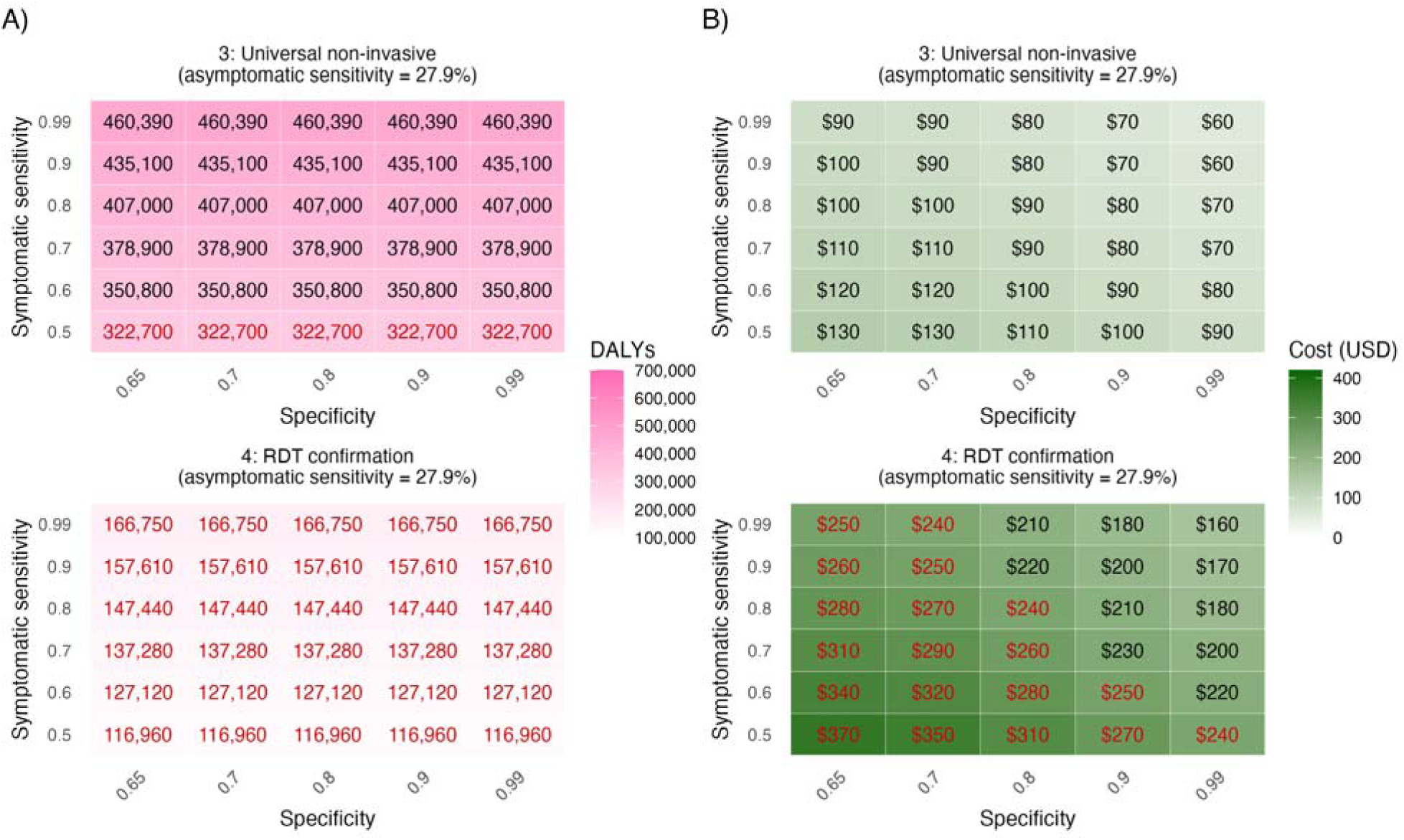
**A)** Disability adjusted life years (DALYs) averted for universal non-invasive diagnostic implementation and non-invasive screening with rapid diagnostic test (RDT) confirmation (strategies 3 and 4) with 27.9% asymptomatic sensitivity. Results are shown for base case scenario. Darker pink tiles indicate greater DALYs averted; red text indicates performance lower than a full RDT scale-up. **B)** Cost per DALY averted for universal non-invasive diagnostic implementation and non-invasive screening with rapid diagnostic test (RDT) confirmation (strategies 3 and 4) with 27.9% asymptomatic sensitivity. Results are shown for base case scenario. Darker green tiles indicate higher cost per DALY averted; red text indicates performance lower than a full RDT scale-up.

Production costs ranged from $50–120 for universal NID implementation when the asymptomatic sensitivity was set to 27.9%, and from $40–80 at 55% (**Figure 4A**). These costs were lower than those associated with full-scale RDT implementation. However, universal NID yielded negative net production costs except when specificity was set to 99% (**Figure 4B**). For NID screening followed by confirmatory RDT testing methods, production costs ranged from $150–410 when the NID asymptomatic sensitivity was 27.9%, and from $120–270 at 55%. Across both asymptomatic sensitivities, this strategy yielded lower production costs than full-scale RDT implementation at higher specificities (**Figure 4A**). In contrast to universal NID implementation, NID screening followed by a confirmatory yielded consistently positive net production costs of $240–2,280 when asymptomatic sensitivity was 27.9% and from $180–810 when asymptomatic sensitivity was 55% (**Figure 4B**).

**Figure 4.**
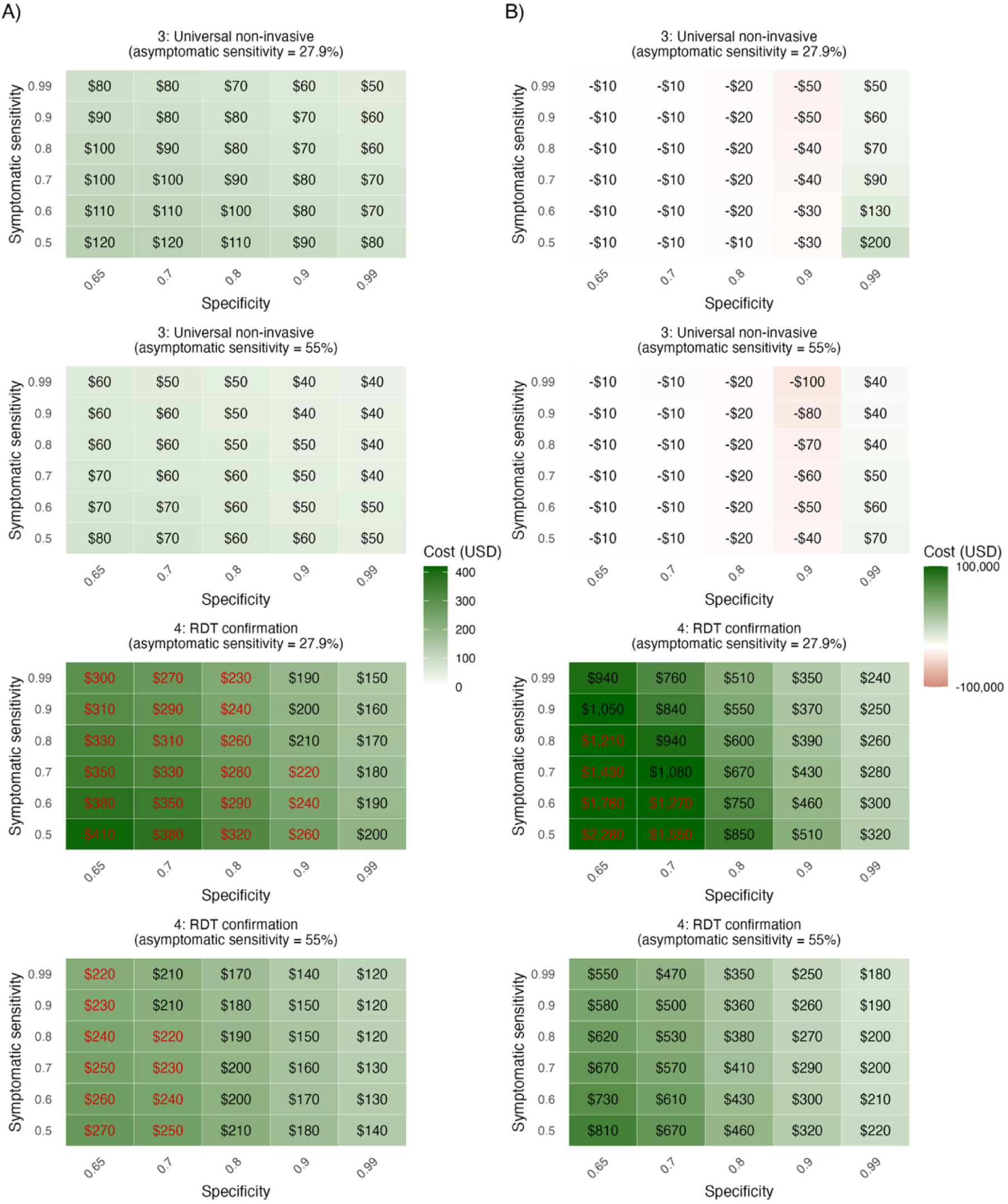
**A)** Production costs for universal non-invasive diagnostic implementation and non-invasive screening with rapid diagnostic test (RDT) confirmation (strategies 3 and 4) with varied asymptomatic sensitivity (27.9% and 55%). Results are shown for base case scenario. Darker green tiles indicate higher production costs; red text indicates performance lower than a full RDT scale-up. **B)** Net production costs for universal non-invasive diagnostic implementation and non-invasive screening with rapid diagnostic test (RDT) confirmation (strategies 3 and 4) with varied asymptomatic sensitivity (27.9% and 55%). Results are shown for base case scenario. Darker green tiles indicate positive cost per net correct diagnosis (true-positive results > excess antimicrobials), while red tiles indicate negative cost per correct diagnosis (excess antimicrobials > true-positive results); red text indicates performance lower than full RDT scale-up.

Taken together, when asymptomatic sensitivity is equal to that of the RDT (27.9%), the NID must attain at least 60% symptomatic sensitivity and above 90% specificity when used alone to achieve improved outcomes across all metrics compared to full-scale RDT use (**Figures 2–4, Figures S9–S13**). When asymptomatic sensitivity is doubled (55%), above 90% specificity is still suggested, mainly to reduce excessive antimicrobial use and net production costs (**Figures 2–4**). When used in combination with RDTs, NIDs do not outperform RDTs alone at any modeled level of test accuracy in terms of case-detection and DALYs averted. However, when used sequentially, they can achieve improved performance across all other metrics (i.e., excess antimicrobials, cost per DALY averted, production cost, and net production cost) with 27.9% asymptomatic sensitivity, a minimum 80% symptomatic sensitivity, and at least 90% specificity.

## Discussion

This study uses a country-agnostic decision-tree model to evaluate the impact of novel NIDs for malaria diagnosis and case detection and aims to define diagnostic accuracy thresholds at which NID adoption is appropriate versus continued RDT investment. While NID research has focused on technical development, few studies have assessed public health impact or provided implementation guidance.^13,26^ Our results suggest NIDs could improve malaria diagnosis across diverse transmission settings compared to syndromic management or standard RDTs, representing a cost-effective intervention to enhance case detection and reduce DALYs. RDTs as a standalone diagnostic method for malaria diagnosis led to improved case detection rates relative to current practice, though full-scale adoption may be financially infeasible. While ICERs of full-scale RDT implementation fall along the cost-effectiveness frontier ($60–1,270/DALY averted at baseline), the strategy required a 36–39% increase in annual budget. Universal NID testing achieved higher detection rates (31–65%) at lower costs through enhanced asymptomatic sensitivity.

The higher asymptomatic sensitivity of NIDs improves all outcomes, given that increased case detection enables timely treatment, which could directly reduce transmission. Additionally, cost savings from NIDs could support broader malaria control efforts. The superior asymptomatic detection and lower unit costs of NIDs relative to RDTs position them as a pivotal tool for scale-up and long-term malaria elimination campaigns. However, standalone NID use generates excess false-positives and inappropriate antimalarial prescriptions, potentially impacting antimalarial supply and enabling resistance development.^41^ Therefore, prioritizing specificity, even at the expense of some sensitivity, could enhance NID acceptability by reducing false-positives and excessive prescribing while still outperforming RDTs. Sequential NID screening with RDT confirmation can also reduce overprescription while lowering costs compared to current practice and universal RDT implementation.

Improvements in NID specificity and asymptomatic sensitivity remain critical, chiefly regarding specificity and asymptomatic sensitivity. At baseline modeled test sensitivities, our threshold analysis suggests NIDs with 76–99% specificity reduce excess antimalarials to yield positive net production costs when used alone, or 65–84% specificity when used together with RDTs (**Figure S8**). Given uncertainty in final NID specifications and performance, we compared its performance to full-scale RDT use under two scenarios: asymptomatic sensitivity equivalent to RDTs (27.9%) and nearly double (55%). Even at 27.9% asymptomatic sensitivity (matching RDTs), NIDs outperformed RDTs in case detection. At 55% asymptomatic sensitivity, NIDs achieved comparable case detection while also supporting antimicrobial stewardship through reduced false-positives and positive net production costs of $180–810 (**Figures 2–4**). Overall, our results suggest that an NID with a minimum 55% asymptomatic sensitivity and at least 84% specificity – especially when followed by a confirmatory RDT – could improve case detection, reduce antimicrobial overuse, and limit costs.

This study has several limitations. First, the novelty of NIDs introduces inherent uncertainty regarding their real-world performance. To address this, we simulated a wide range of diagnostic sensitivities and specificities across 36 unique scenarios of varying prevalence and symptomatic proportion. Therefore, these findings are applicable across diverse epidemiological contexts and could guide NID implementation globally. Furthermore, as this analysis evaluates non-invasive malaria diagnostics as a broad class, the conclusions are not limited to a single device. While this approach necessitates underlying assumptions regarding technological heterogeneity, our sensitivity analyses were designed to capture this variability, particularly as many NIDs remain in early development. This study does not use a transmission model, so the impact of reducing asymptomatic infections cannot be fully assessed. NIDs could support elimination by detecting subclinical infections, but dynamic transmission modeling is needed to define the performance thresholds and strategies required for this. As a result, our findings may underestimate the long-term benefits of lowering parasite transmission.

Additionally, this model assumes a theoretical population with universal access to care; consequently, our findings likely overrepresent the clinical impact and cost savings of the proposed strategies, especially given that such large-scale implementation of active case-finding is likely not feasible. Nevertheless, these estimates provide a foundational understanding of the relative advantages offered by NIDs in malaria diagnosis, their relationships to epidemiological conditions, and the general improvement trends they may offer. The scalability of non-invasive technology depends on manufacturing capacity, supply chain logistics, and integration into existing health systems.^42^ Future research should evaluate NID impact across specific age groups and high-priority settings, including schools, high-burden areas, and elimination zones, as field data become available. The cost-effectiveness of RDTs compared to microscopy has been investigated in elimination settings, showing greatest effect but also the highest costs.^43^ Appropriately priced NIDs could potentially address this gap, as rapid and accurate screening tools remain critical for enhancing surveillance and accelerating elimination.

There have been several studies that have evaluated the acceptability and feasibility of non-invasive diagnostics using urine or saliva samples, suggesting that scaling-up these non-invasive technologies point-of-care implementation is possible with limited laboratory infrastructure.^16,17,19^ Near-infrared spectroscopy (NIRS) and volatile organic compound detectors are also emerging as viable NIDs.^12,15,18^ While their operational feasibility remains to be fully established, stakeholders in Peru have expressed a preference for these NIDs, which may offer additional distinct advantages in settings where non-invasive tools like saliva tests are already accepted by eliminating the need for sample collection and reducing consumable waste.^14,44,45^ Specific NID-related maintenance fees and additional consumables were omitted from our cost analysis due to substantial variation across NID platforms. Thus, our results may overestimate cost savings again by not accounting for the unforeseen operational expenses associated with field-level deployment or alternatively, underestimate cost savings if instruments are developed at lower price points. Future cost analyses are needed to determine appropriate price points of NIDs and their use over time.

As the World Health Organization ultimately aims to reduce malaria incidence and mortality rates by at least 90% by the year 2030, the integration of non-invasive technology in diagnostic pathways, whether used alone or paired with RDTs, presents as a promising strategy to enhance malaria detection and clinical management.^46^ Our findings indicate that pairing NIDs with RDTs can maintain timely case detection while substantially reducing inappropriate antimicrobial use, thereby improving clinical management and antimicrobial stewardship. By reserving confirmatory RDTs for NID-positive individuals, this approach enables more efficient use of constrained health system resources and lowers overall costs. Taken together, these results suggest that non-invasive diagnostic technologies could play a meaningful role in strengthening malaria control strategies, particularly as preserving therapeutic efficacy becomes increasingly critical in the face of rising antimicrobial resistance.

## Data Availability

We acknowledge the importance of transparent and responsible data sharing to foster scientific collaboration and advance research. The publicly available data used in the study do not require de-identification. All necessary data is included in the manuscript and supplementary materials. For additional information, interested parties can contact MAH.

## Declarations

### Ethics approval and consent to participate

This modeling study used publicly available data and did not require formal ethical approval. Ethical considerations were integrated throughout, and the methodology is fully documented in the manuscript and Table 1. We declare no conflicts of interest, reflecting our commitment to ethical standards.

### Consent for publication

Not applicable.

### Authors’ contributions

Conceptualization: MAH, KKAT, BEN. Data curation: MAH, BEN. Methodology: MAH, AdN, BEN. Formal analysis: MAH, AdN. Investigation: MAH. Supervision: BEN. Writing – original draft: MAH, SC, BEN. Writing – review and editing: MAH, AdN, SC, KKAT, BEN.

## Acknowledgements

Not applicable.

## Supplementary Appendix

**Text S1.**
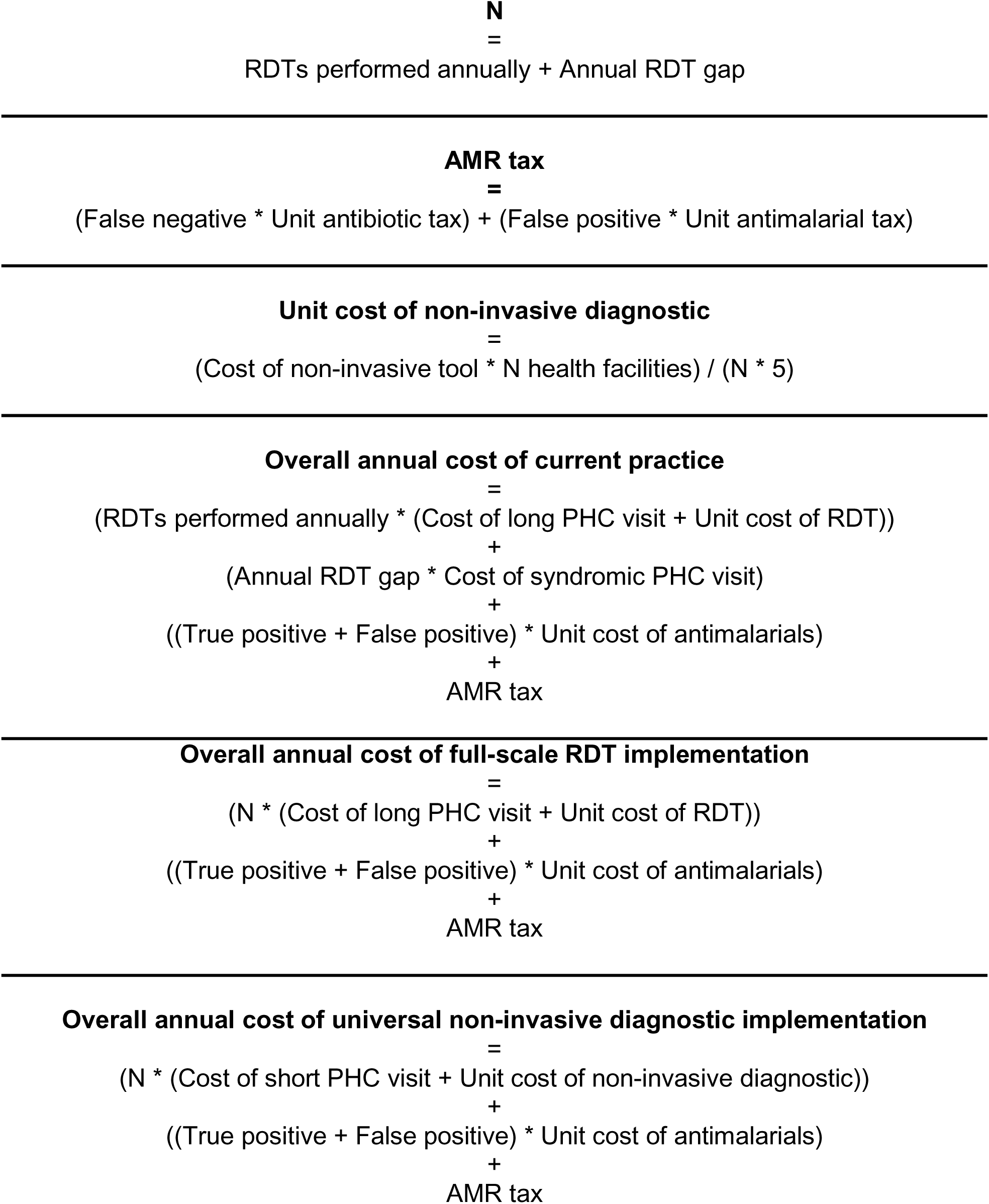
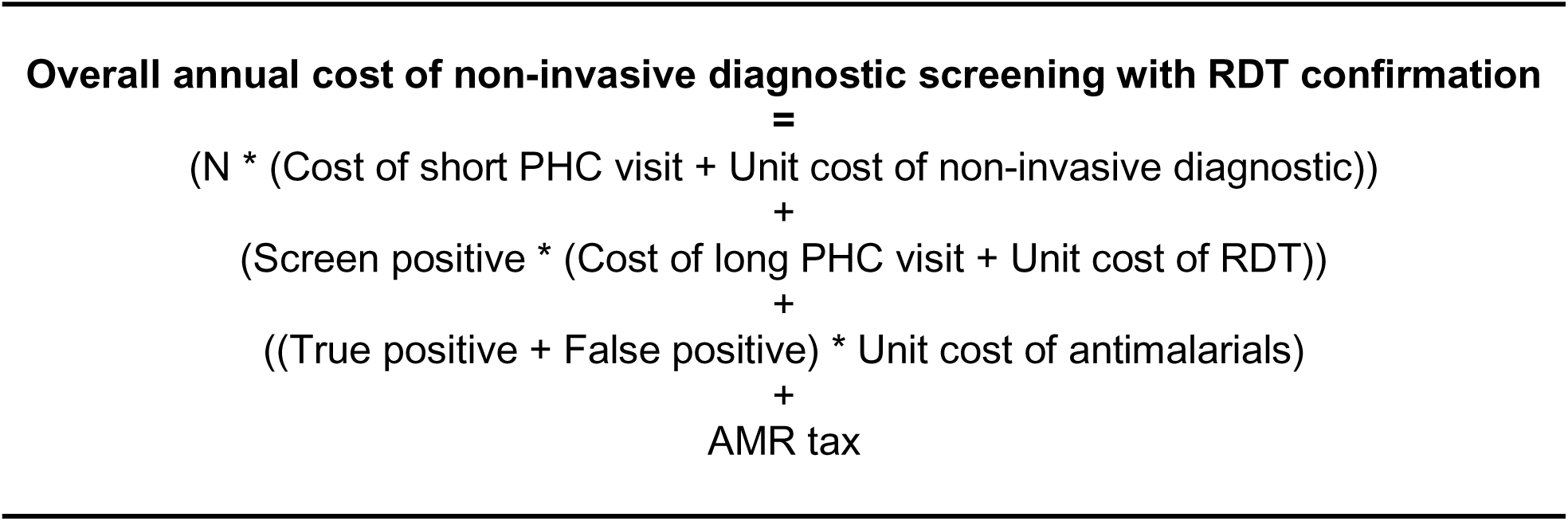
Explanation of cost calculations. The calculations for antimicrobial resistance (AMR) taxes, unit cost of the non-invasive diagnostic, and overall annual cost of each testing strategy are outlined below.

**Figure S1.**
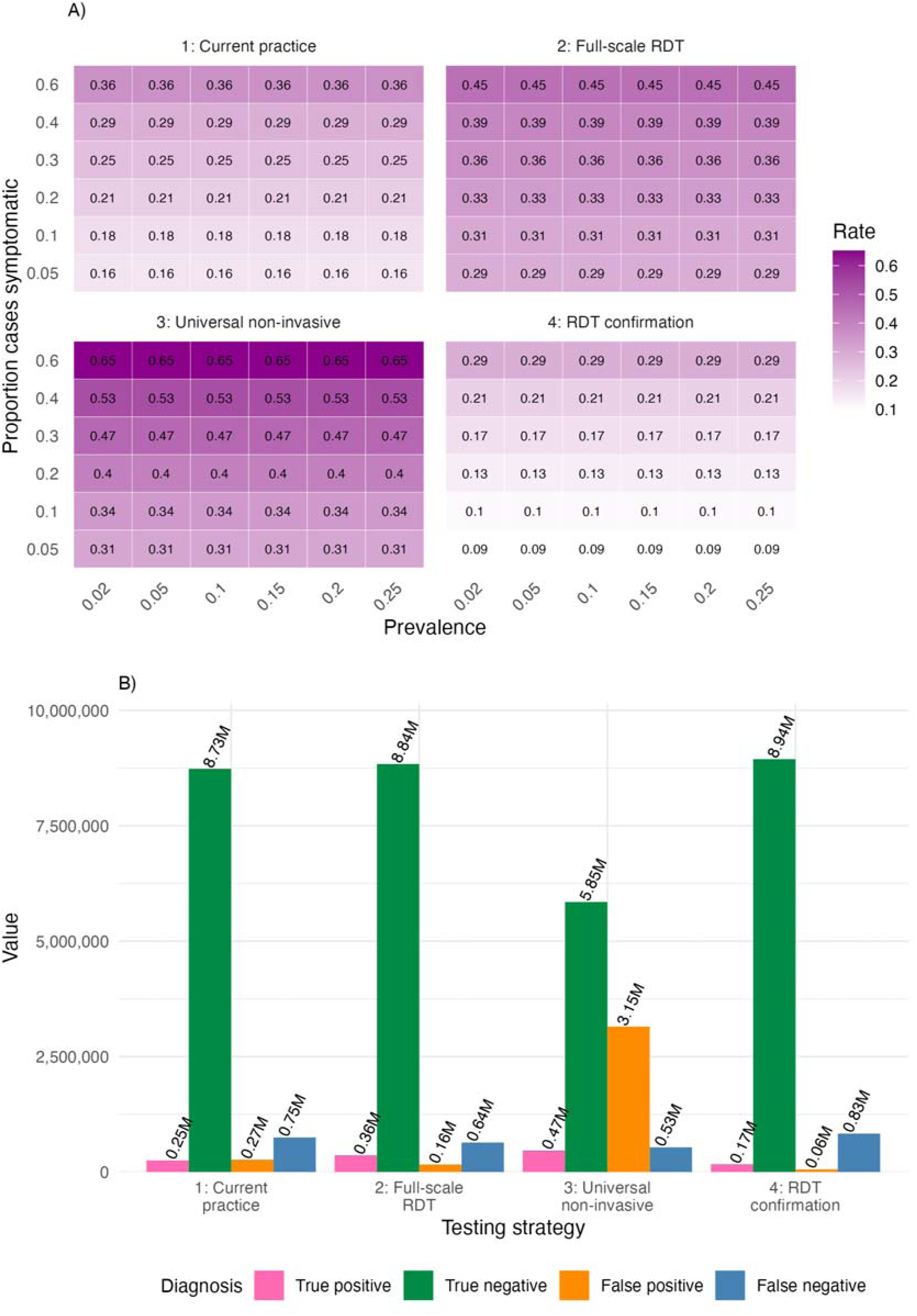
**A)** Overall case detection rate for each testing strategy across varied malaria prevalence values and symptomatic proportion at the base case (i.e., 10% malaria prevalence 30% symptomatic proportion). Darker violet tiles indicate higher case detection rates. **B)** Diagnosis status by testing strategy. Results shown are for baseline NID accuracy (Table 1).

**Figure S2.**
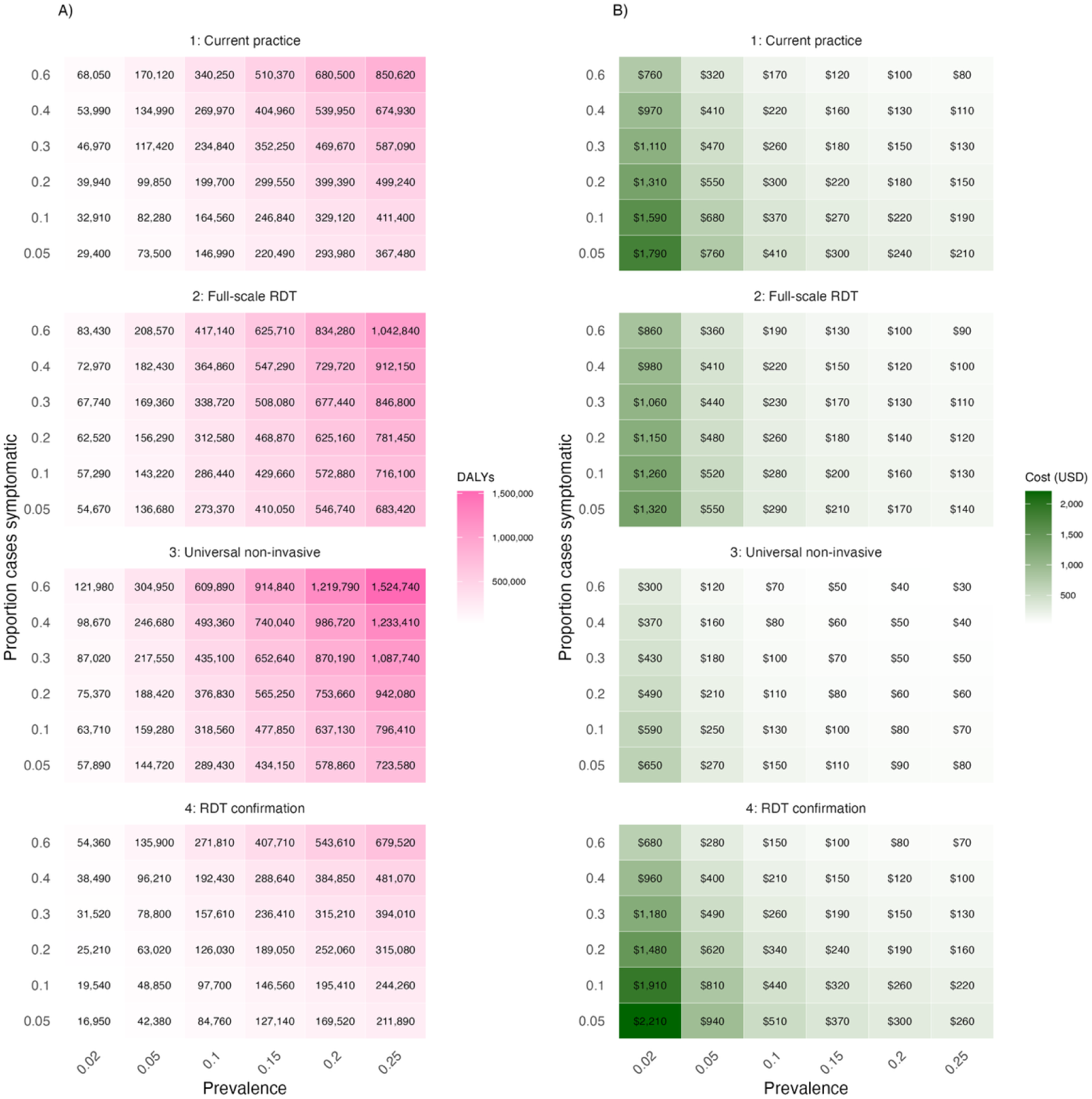
**A)** Disability adjusted life years (DALYs) averted for each testing strategy across varied malaria prevalence values and symptomatic proportion. Darker violet tiles indicate greater DALYs averted. **B)** Cost per DALYs averted for each testing strategy across varied malaria prevalence values and symptomatic proportion. Darker green tiles indicate greater cost per DALY averted. Results shown are for baseline NID accuracy (Table 1).

**Figure S3.**
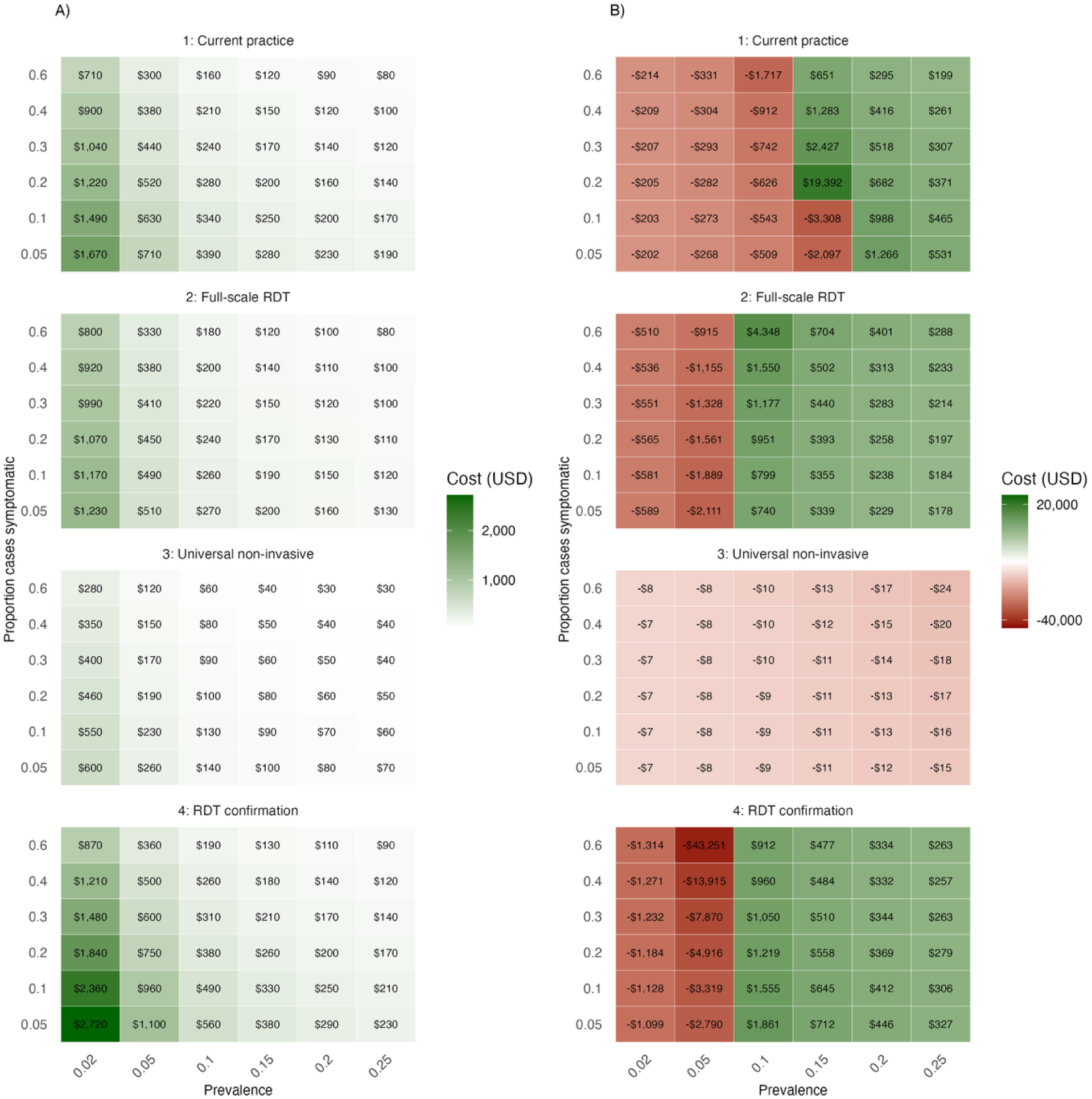
**A)** Production cost for each testing strategy across varied malaria prevalence values and symptomatic proportion. Darker green tiles indicate higher production costs. **B)** Cost per net correct diagnosis for each testing strategy across varied malaria prevalence values and symptomatic proportion. Darker green tiles indicate positive cost per net correct diagnosis (i.e., more true-positive results than excess antimicrobials), while red tiles indicate negative cost per correct diagnosis (i.e., more excess antimicrobials than true-positive results). Results shown are for baseline NID accuracy (Table 1).

**Figure S4.**
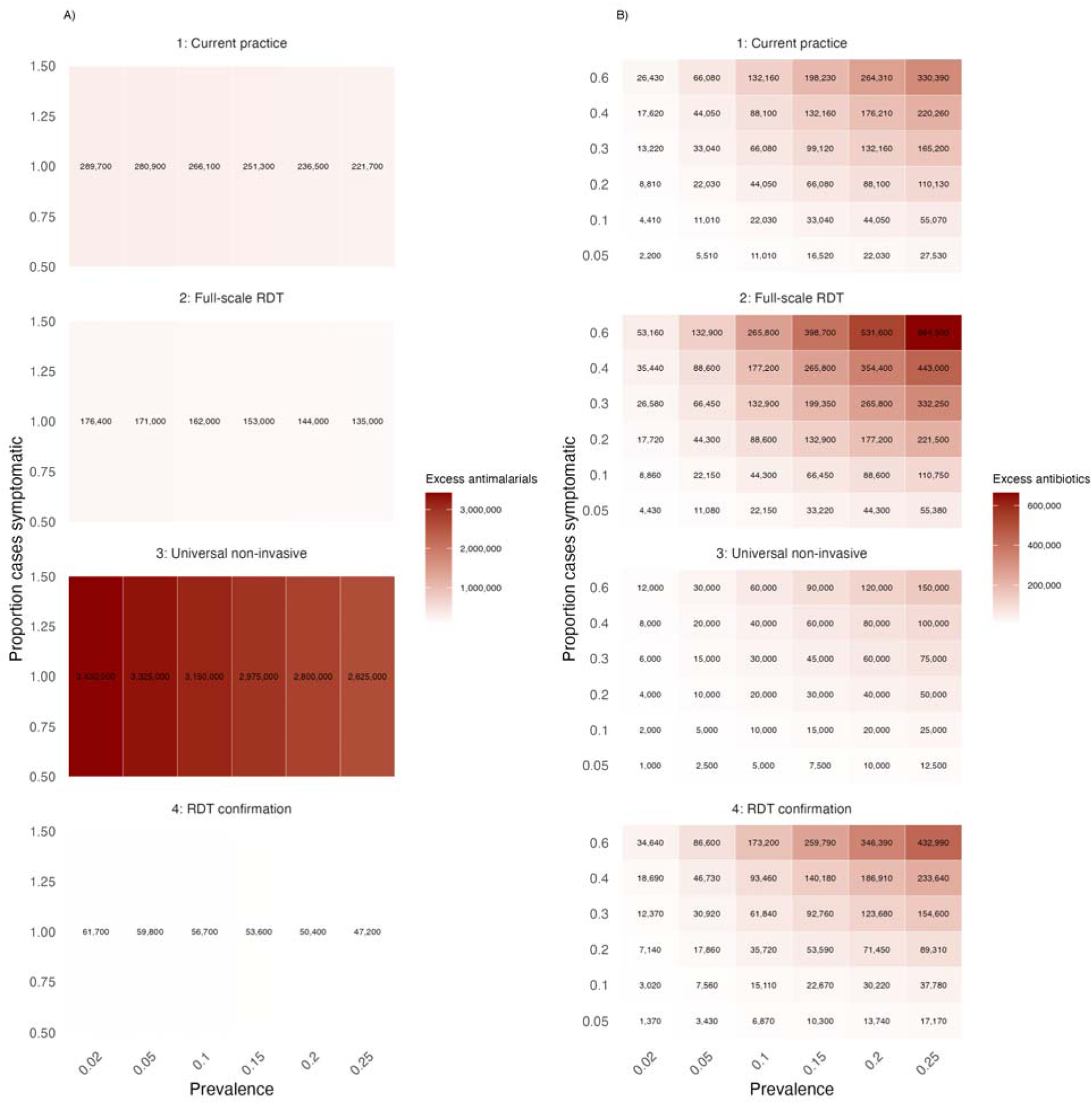
**A)** Excess antimalarials prescribed for each testing strategy across varied malaria prevalence values and symptomatic proportion. Darker red tiles indicate greater values of excess antimalarials. **B)** Excess antibiotics prescribed for each testing strategy across varied malaria prevalence values and symptomatic proportion. Darker red tiles indicate greater values of excess antibiotics. Results shown are for baseline NID accuracy (Table 1).

**Figure S5.**
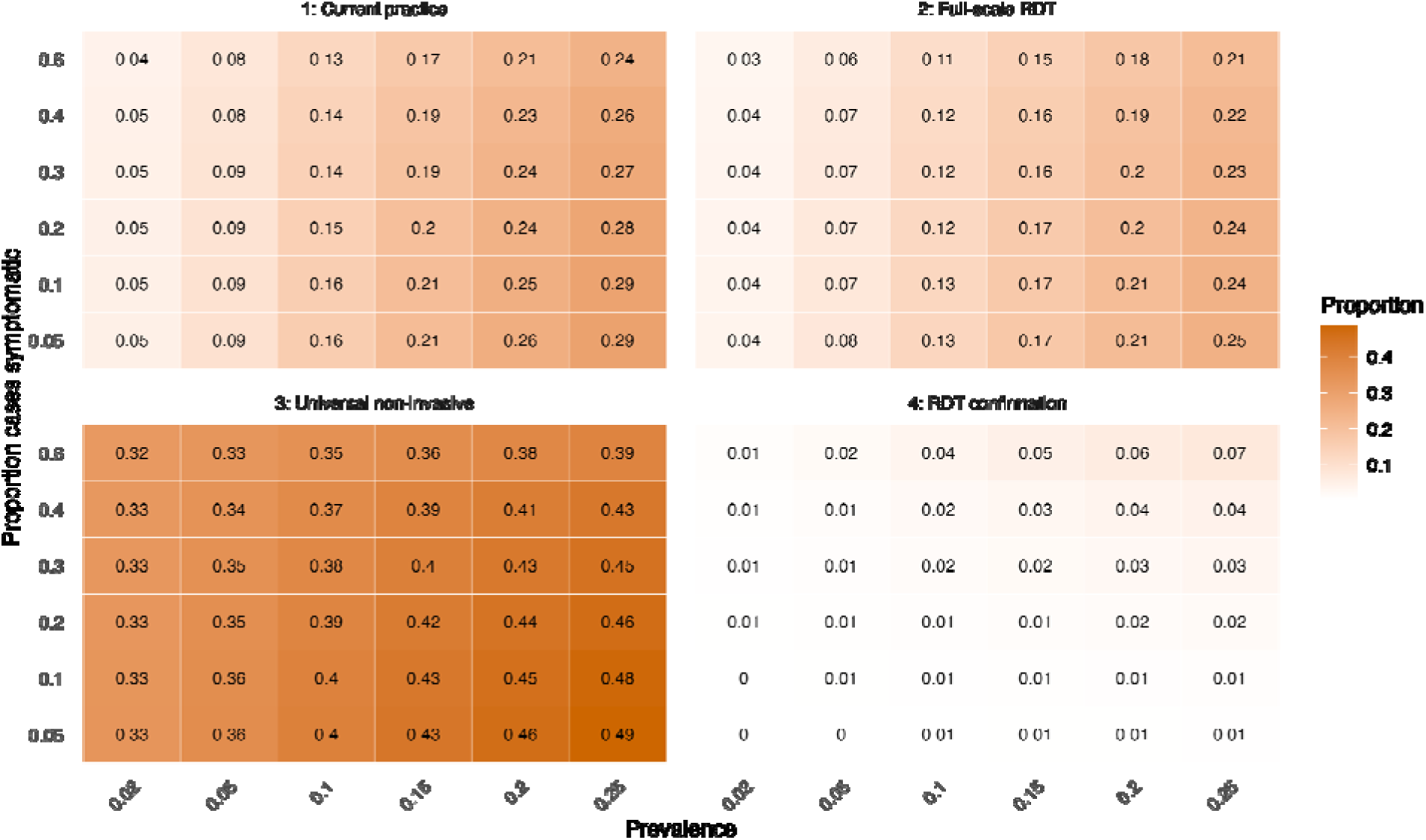
Proportion of overall costs attributable to unnecessary prescriptions (i.e., excess antimalarials and antibiotics) for each testing strategy across varied malaria prevalence values and symptomatic proportion. Darker orange tiles indicate higher proportions of cost from excess prescriptions. Results shown are for baseline NID accuracy (Table 1).

**Figure S6.**
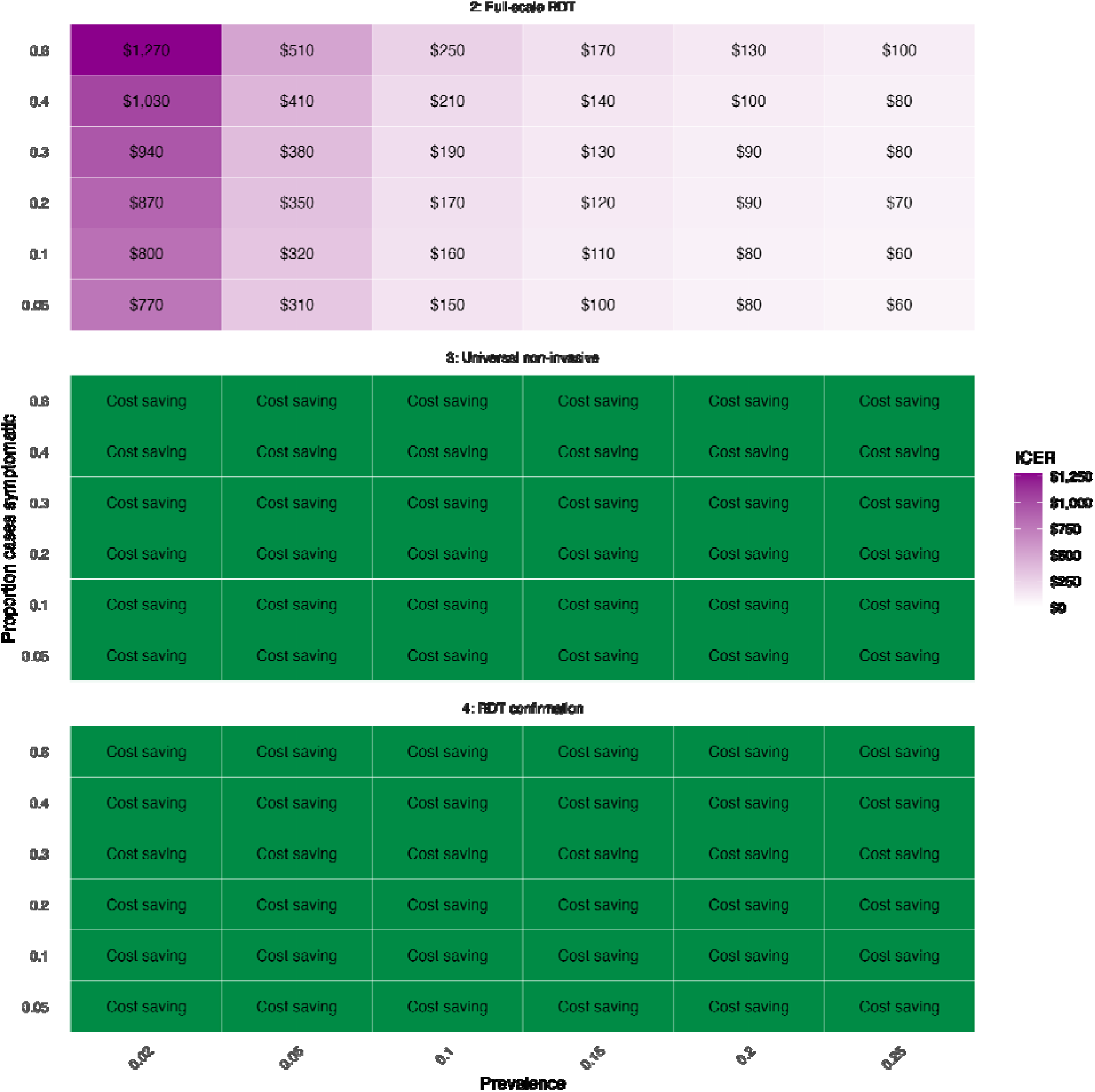
Results from 36 distinct cost-effectiveness analyses within each varied malaria prevalence value and symptomatic proportions comparing each testing strategy to the baseline strategy (current practice). Green tiles represent cost saving scenarios. Darker violet tiles represent greater ICER values. Results shown are for baseline NID accuracy (Table 1).

**Figure S7.**
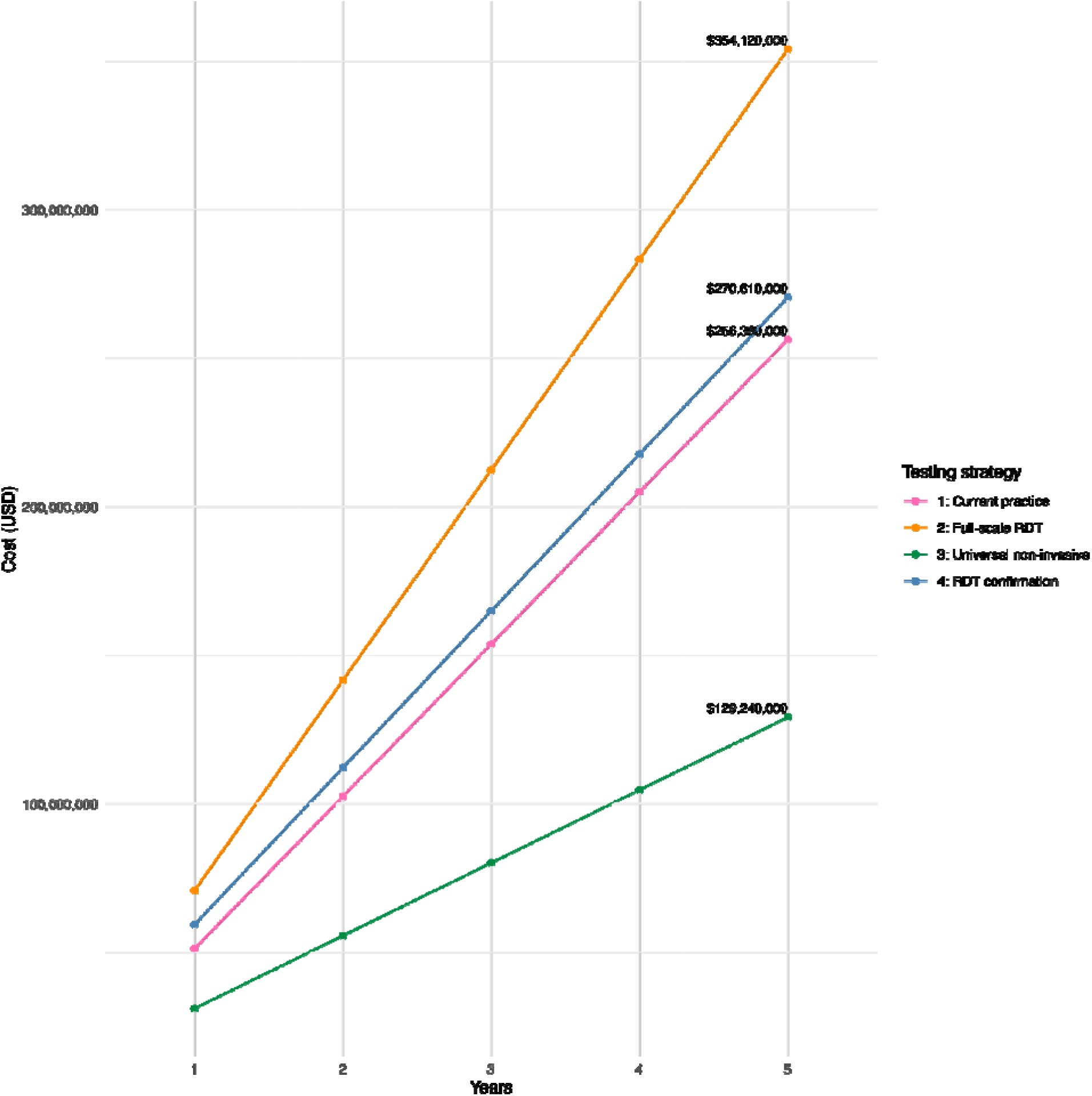
Long-term overall costs (5 years) by testing strategy. Results shown are for baseline NID accuracy (Table 1) and base case epidemic conditions: 10% malaria prevalence and 30% symptomatic proportion.

**Figure S8.**
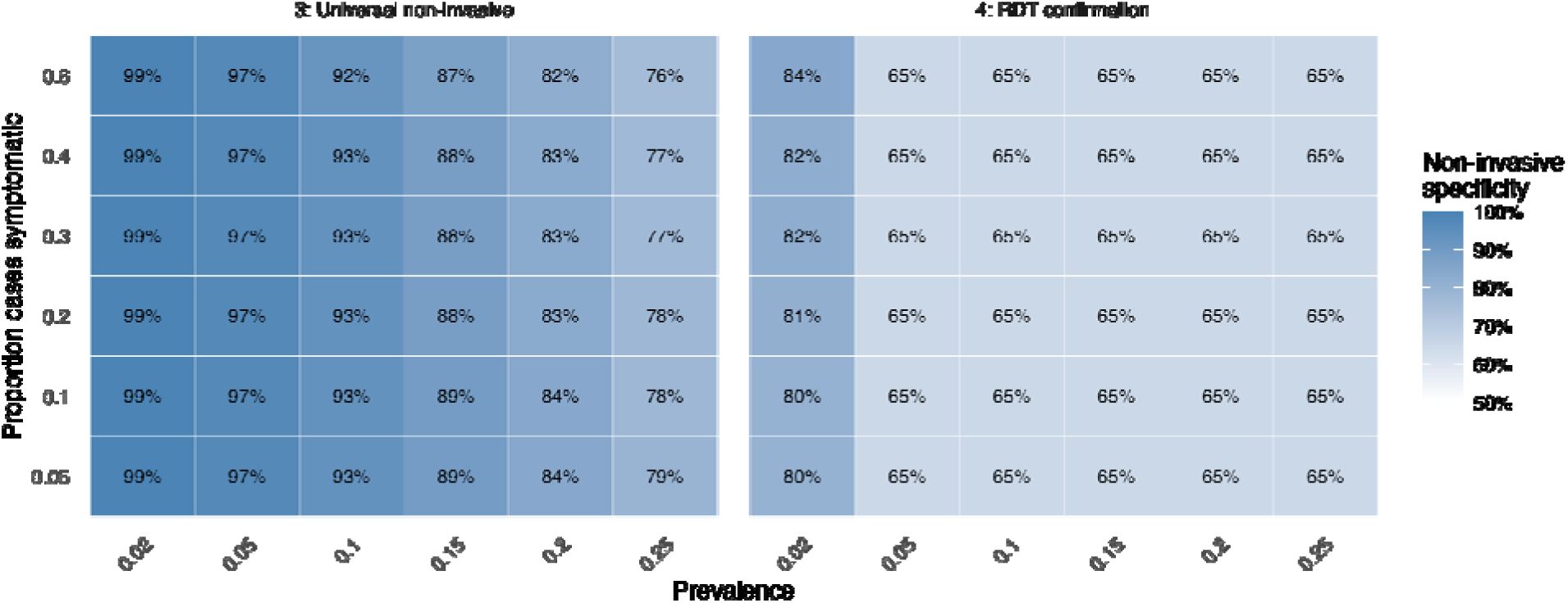
Threshold analysis results representing the minimum non-invasive diagnostic specificity required to have a positive net cost per correct diagnosis across varied malaria prevalence values and symptomatic proportion. Darker blue tiles indicate higher required specificity.

**Figure S9.**
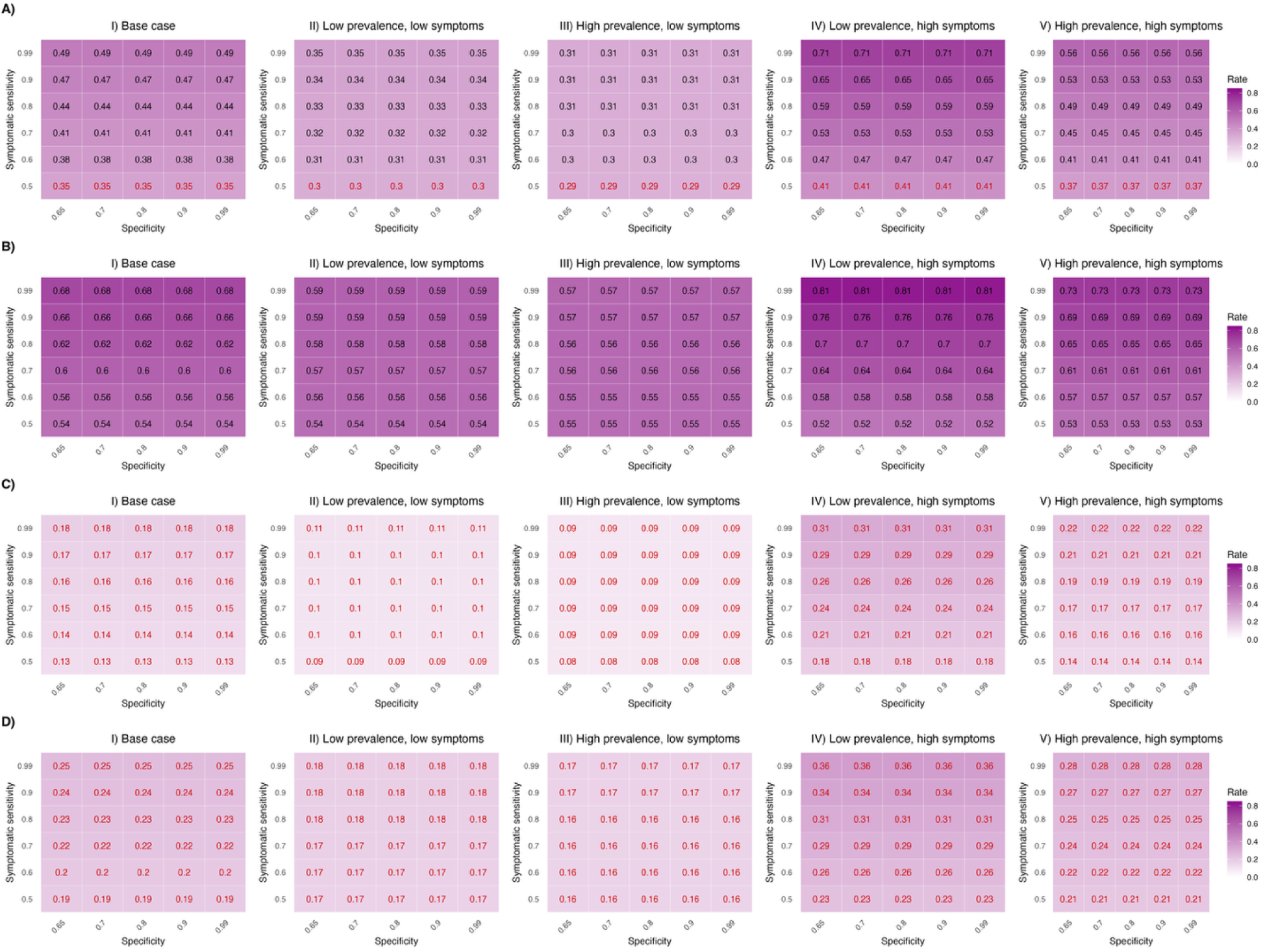
Case detection rates for universal non-invasive diagnostic implementation and non-invasive screening with rapid diagnostic test (RDT) confirmation (strategies 3 and 4) under five epidemic conditions. **Panels A and B** correspond to universal non-invasive diagnostic implementation, with asymptomatic sensitivity equal to that of an RDT (27.9%) and twice that value (55%), respectively. **Panels C and D** show results for non-invasive screening with RDT confirmation using the same sensitivity assumptions. Darker violet tiles indicate higher case detection rates; red text indicates performance lower than a full RDT scale-up. Epidemic conditions are: **I)** base case (10% prevalence, 30% symptomatic), **II)** low prevalence & low symptoms (2%, 10%), **III)** high prevalence & low symptoms (20%, 5%), **IV)** low prevalence & high symptoms (5%, 60%), and **V)** high prevalence & high symptoms (25%, 40%).

**Figure S10.**
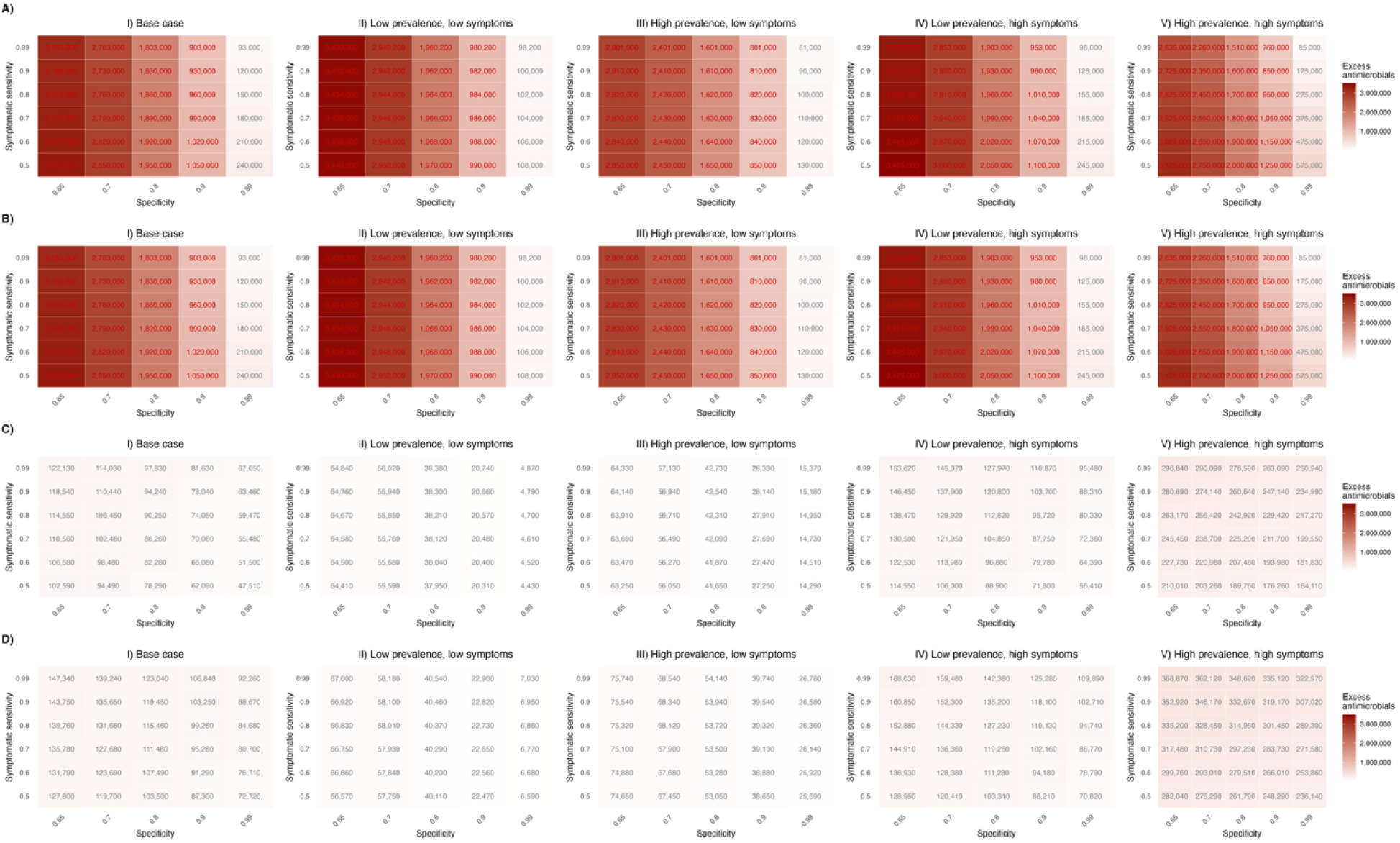
Excess prescriptions (antimalarials and antimicrobials) averted for universal non-invasive diagnostic implementation and non-invasive screening with rapid diagnostic test (RDT) confirmation (strategies 3 and 4) under five epidemic conditions. **Panels A and B** correspond to universal non-invasive diagnostic implementation, with asymptomatic sensitivity equal to that of an RDT (27.9%) and twice that value (55%), respectively. **Panels C and D** show results for non-invasive screening with RDT confirmation using the same sensitivity assumptions. Darker red tiles indicate greater excess prescriptions; red text indicates performance lower than a full RDT scale-up. Epidemic conditions are: **I)** base case (10% prevalence, 30% symptomatic), **II)** low prevalence & low symptoms (2%, 10%), **III)** high prevalence & low symptoms (20%, 5%), **IV)** low prevalence & high symptoms (5%, 60%), and **V)** high prevalence & high symptoms (25%, 40%).

**Figure S11.**
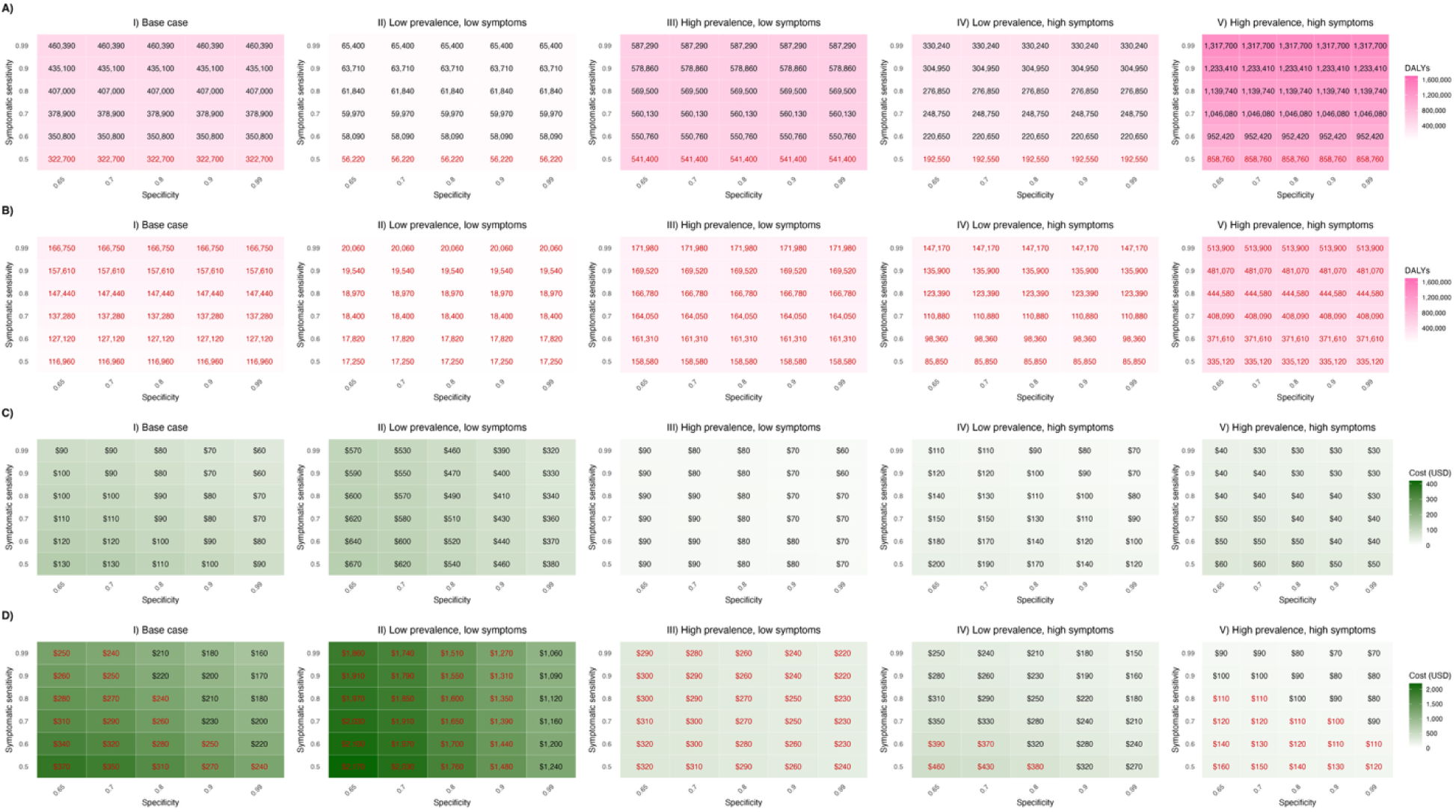
**Panels A and B** display disability adjusted life years (DALYs) averted for universal non-invasive diagnostic implementation and non-invasive screening with rapid diagnostic test (RDT) confirmation (strategies 3 and 4) respectively under five epidemic conditions with NID asymptomatic sensitivity equal to that of an RDT (27.9%). Darker pink tiles indicate greater DALYs averted; red text indicates performance lower than a full RDT scale-up. **Panels C and D** display cost per DALY averted for universal non-invasive diagnostic implementation and non-invasive screening with RDT confirmation (strategies 3 and 4) respectively under five epidemic conditions with the NID asymptomatic sensitivity equal to that of an RDT (27.9%). Darker green tiles indicate higher costs per DALY averted; red text indicates performance lower than a full RDT scale-up. Epidemic conditions are: **I)** base case (10% prevalence, 30% symptomatic), **II)** low prevalence & low symptoms (2%, 10%), **III)** high prevalence & low symptoms (20%, 5%), **IV)** low prevalence & high symptoms (5%, 60%), and **V)** high prevalence & high symptoms (25%, 40%).

**Figure S12.**
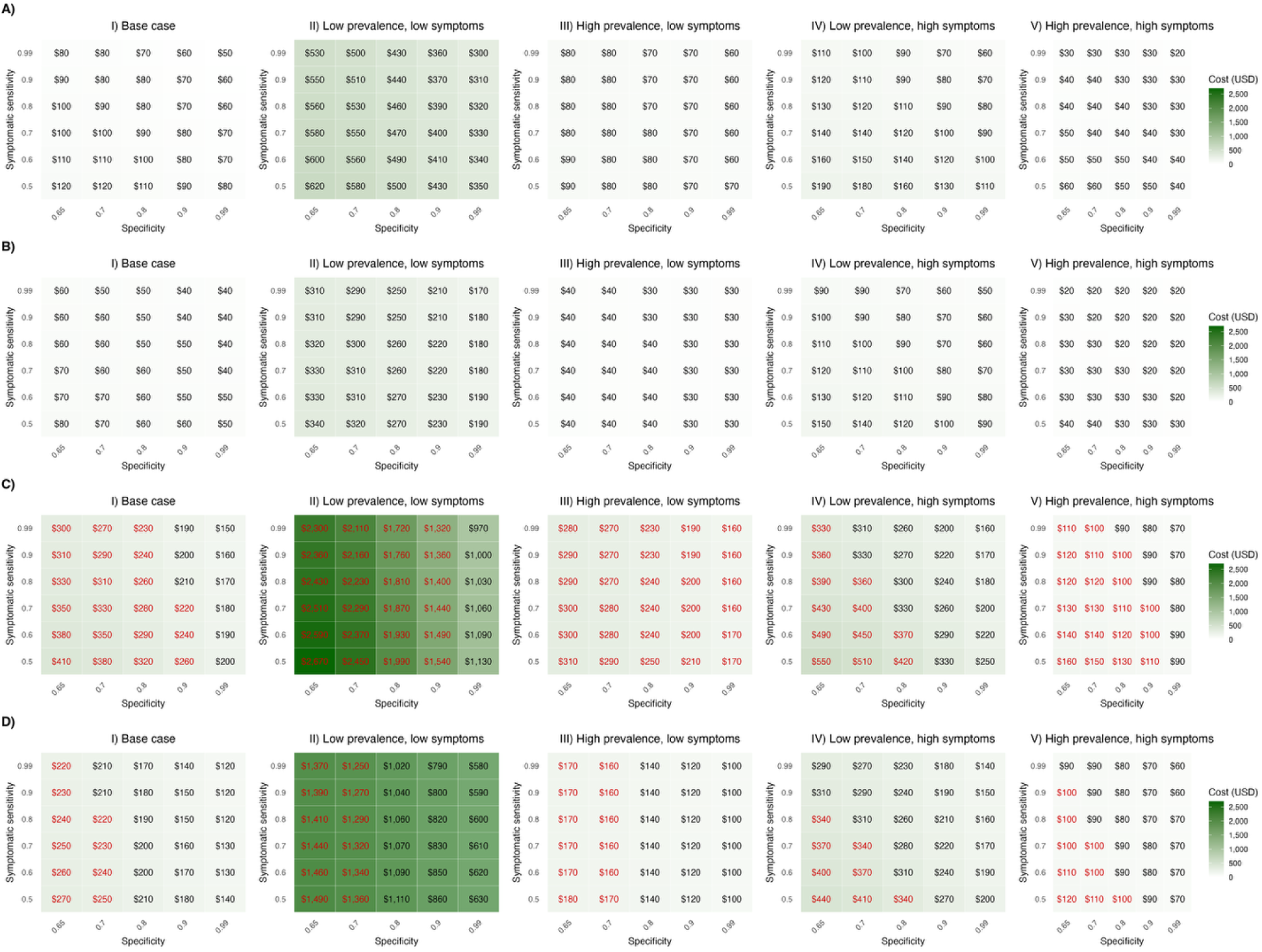
Production cost for universal non-invasive diagnostic implementation and non-invasive screening with rapid diagnostic test (RDT) confirmation (strategies 3 and 4) under five epidemic conditions. **Panels A and B** correspond to universal non-invasive diagnostic implementation, with asymptomatic sensitivity equal to that of an RDT (27.9%) and twice that value (55%), respectively. **Panels C and D** show results for non-invasive screening with RDT confirmation using the same sensitivity assumptions. Darker green tiles indicate higher production costs; red text indicates performance lower than a full RDT scale-up. Epidemic conditions are: **I)** base case (10% prevalence, 30% symptomatic), **II)** low prevalence & low symptoms (2%, 10%), **III)** high prevalence & low symptoms (20%, 5%), **IV)** low prevalence & high symptoms (5%, 60%), and **V)** high prevalence & high symptoms (25%, 40%).

**Figure S13.**
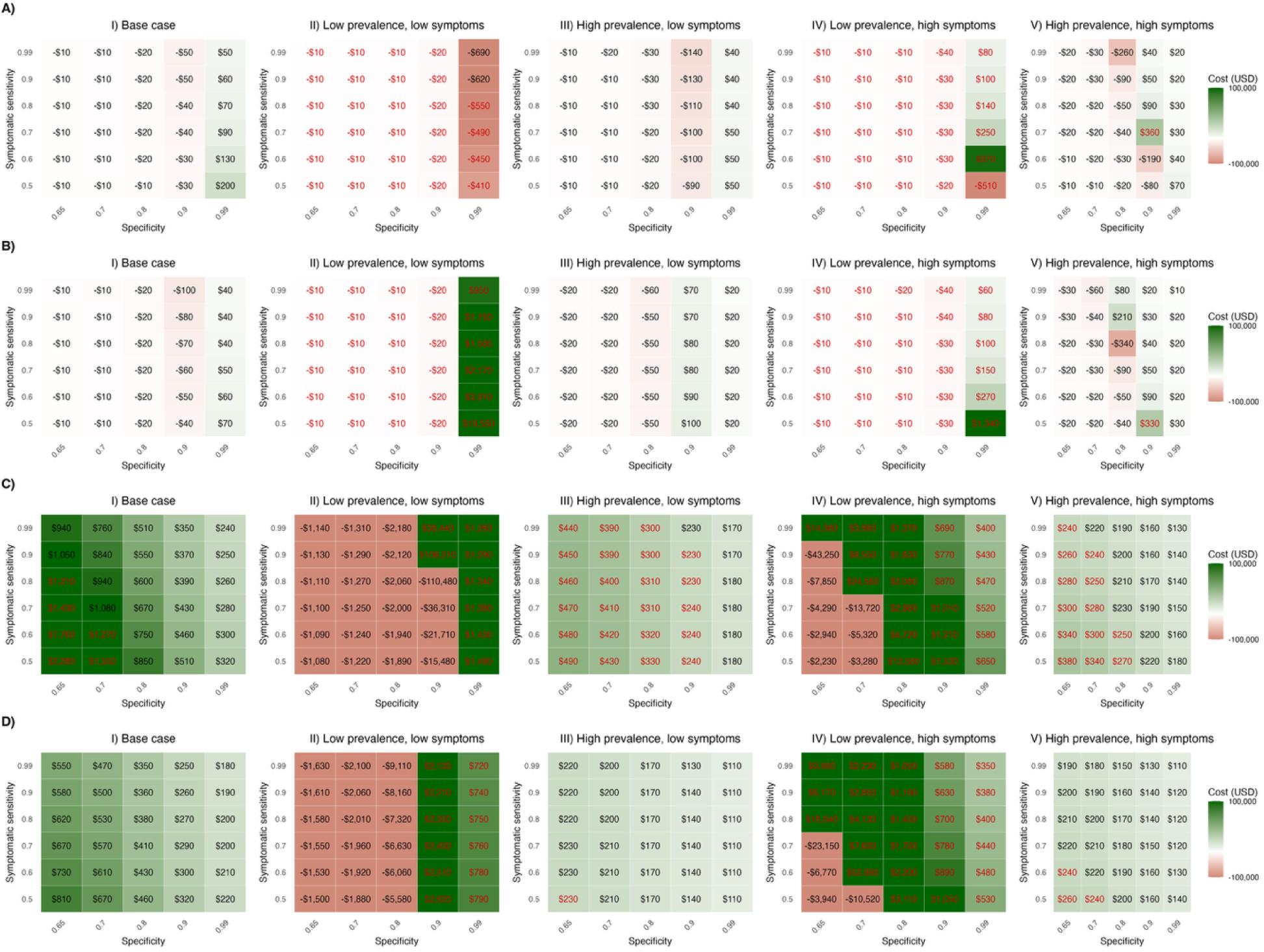
Net production cost for universal non-invasive diagnostic implementation and non-invasive screening with rapid diagnostic test (RDT) confirmation (strategies 3 and 4) under five epidemic conditions. **Panels A and B** correspond to universal non-invasive diagnostic implementation, with asymptomatic sensitivity equal to that of an RDT (27.9%) and twice that value (55%), respectively. **Panels C and D** show results for non-invasive screening with RDT confirmation using the same sensitivity assumptions. Darker green tiles indicate positive cost per net correct diagnosis (more true-positive results than excess antimicrobials), while red tiles indicate negative cost per correct diagnosis (more excess antimicrobials than true-positive results); red text indicates performance lower than a full RDT scale-up. Epidemic conditions are: **I)** base case (10% prevalence, 30% symptomatic), **II)** low prevalence & low symptoms (2%, 10%), **III)** high prevalence & low symptoms (20%, 5%), **IV)** low prevalence & high symptoms (5%, 60%), and **V)** high prevalence & high symptoms (25%, 40%).

**Table S1.**
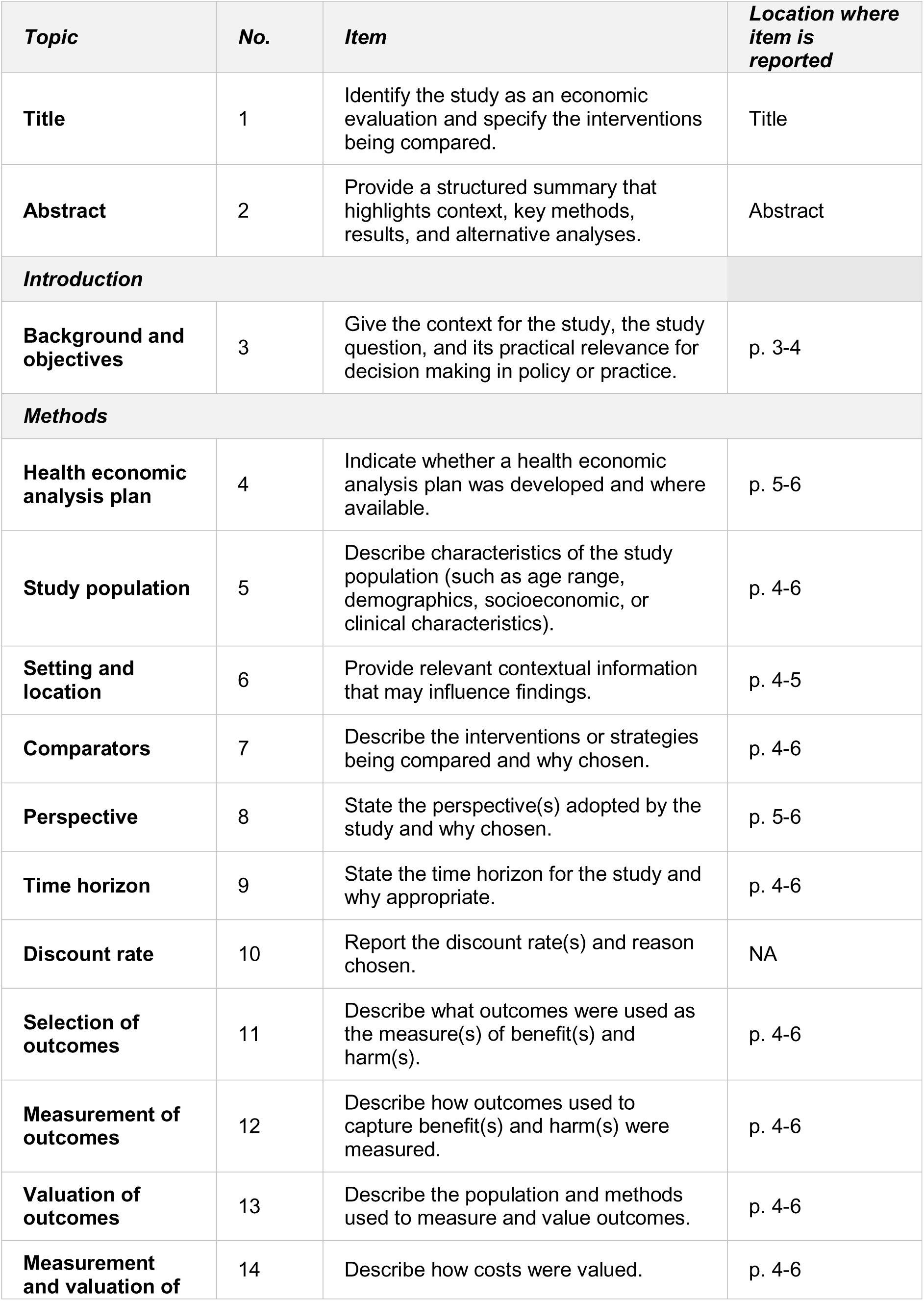
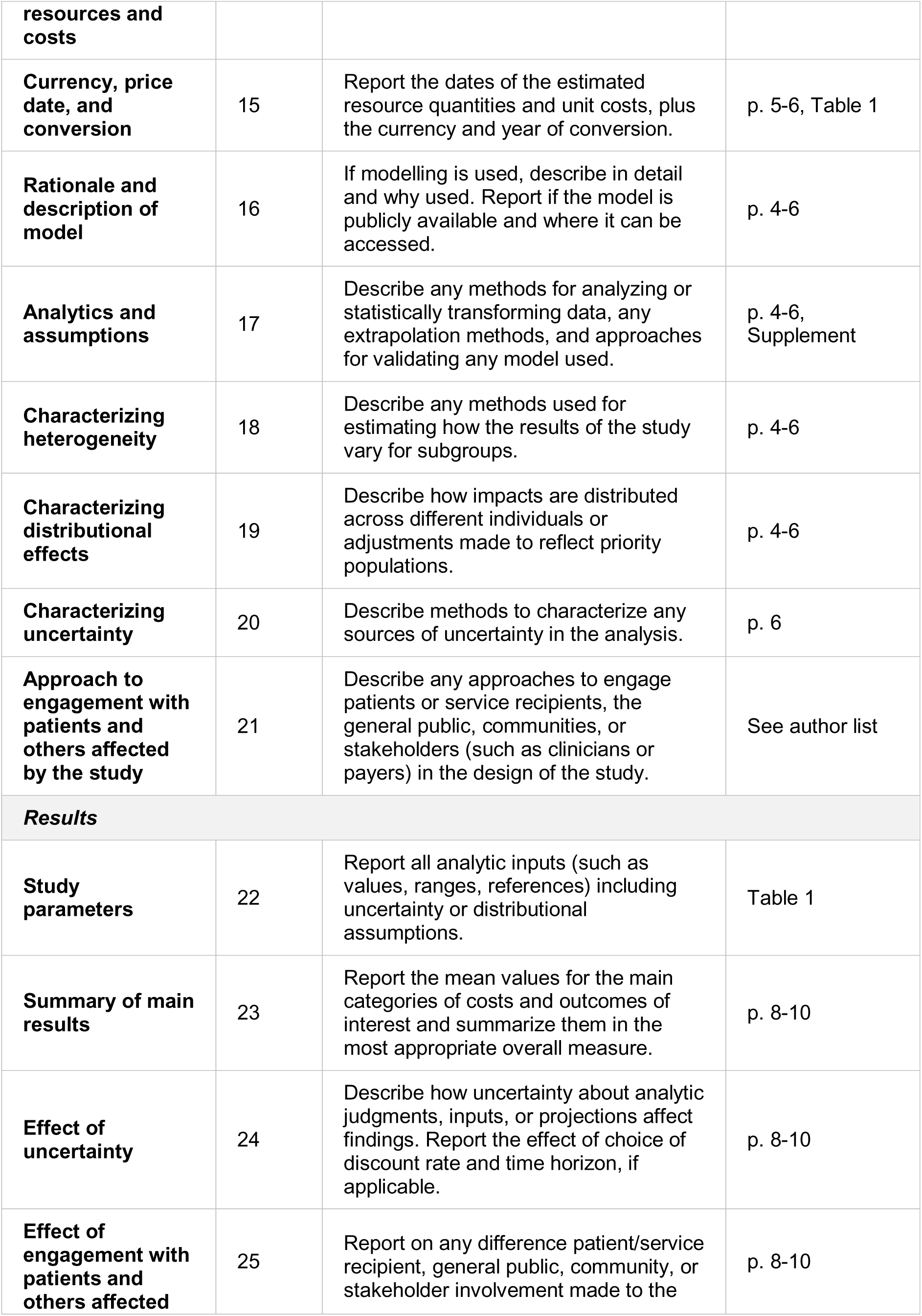
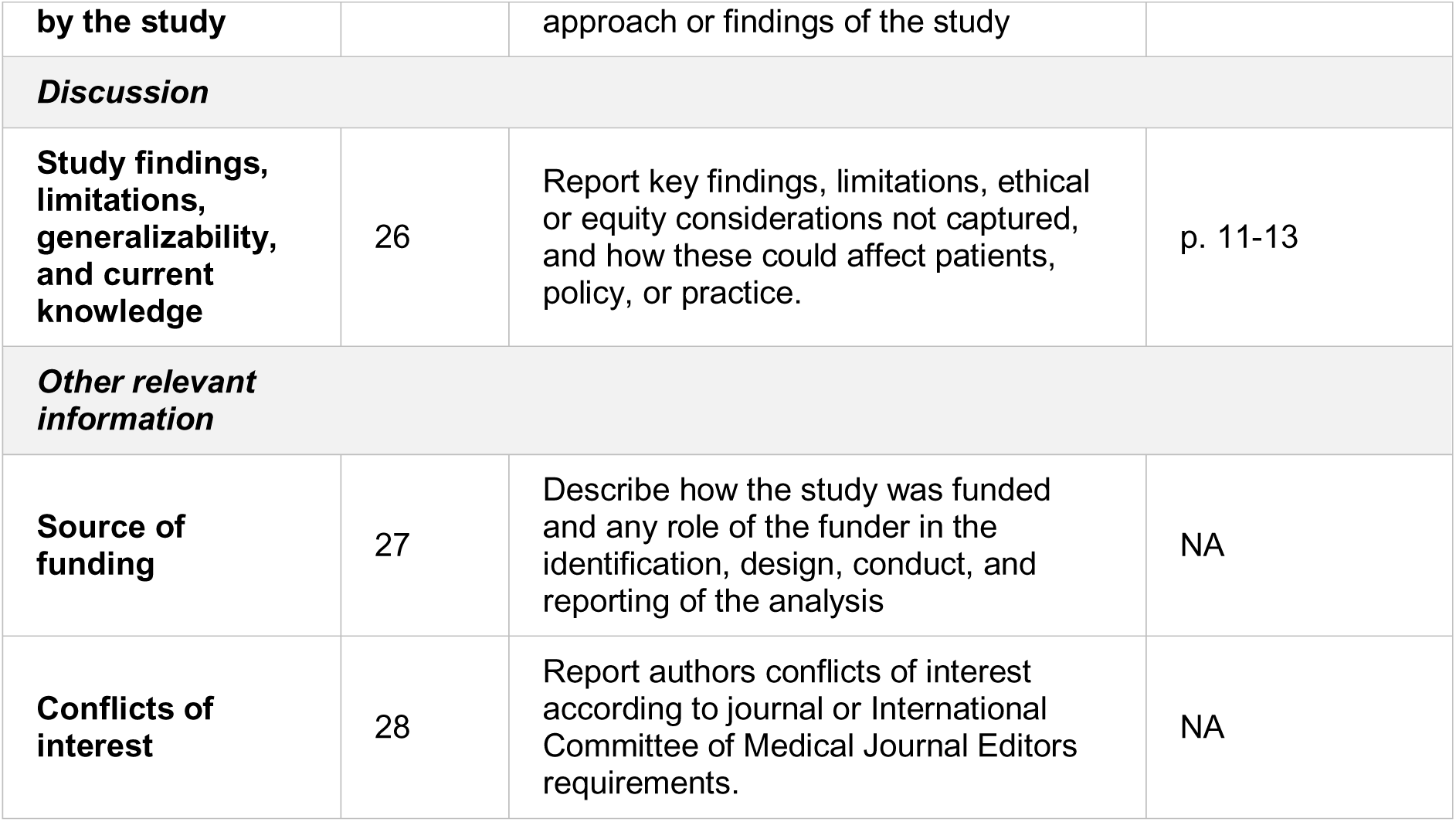
CHEERS checklist.

